# SARS-CoV-2 specific T cells and antibodies in COVID-19 protection: a prospective study

**DOI:** 10.1101/2021.08.19.21262278

**Authors:** Ivan A. Molodtsov, Evgenii Kegeles, Alexander N. Mitin, Olga Mityaeva, Oksana E. Musatova, Anna E. Panova, Mikhail V. Pashenkov, Iuliia O. Peshkova, Alsalloum Almaqdad, Walaa Asaad, Anna S. Budikhina, Aleksander S. Deryabin, Inna V. Dolzhikova, Ioanna N. Filimonova, Alexandra N. Gracheva, Oxana I. Ivanova, Anastasia Kizilova, Viktoria V. Komogorova, Anastasia Komova, Natalia I. Kompantseva, Denis A. Lagutkin, Yakov A. Lomakin, Alexandra V. Maleeva, Elena V. Maryukhnich, Afraa Mohammad, Vladimir V. Murugin, Nina E. Murugina, Anna Navoikova, Margarita F. Nikonova, Leyla A. Ovchinnikova, Natalia V. Pinegina, Daria M. Potashnikova, Elizaveta V. Romanova, Aleena A. Saidova, Nawar Sakr, Anastasia G. Samoilova, Yana Serdyuk, Naina T. Shakirova, Nina I. Sharova, Savely A. Sheetikov, Anastasia F. Shemetova, Liudmila Shevkova, Alexander V. Shpektor, Anna Trufanova, Anna V. Tvorogova, Valeria M. Ukrainskaya, Anatoliy S. Vinokurov, Daria A. Vorobyeva, Ksenia V. Zornikova, Grigory A. Efimov, Musa R. Khaitov, Ilya A. Kofiadi, Alexey A. Komissarov, Denis Y. Logunov, Nelli B. Naigovzina, Yury P. Rubtsov, Irina A. Vasilyeva, Pavel Volchkov, Elena Vasilieva

## Abstract

Rapid spread of COVID-19 pandemic made a substantial share of the world population immunised by SARS-CoV-2 antigens. Infection induces the development of virus-specific antibodies and T cells. Ample evidence on the antibody-mediated protection is contrasted by the elusive role of T cells in preventing infection. To explore the impact of T cells and to quantify the protective levels of the immune responses we conducted a large prospective study: 5,340 Moscow residents were evaluated for the antibody and cellular immune responses to SARS-CoV-2 and monitored for COVID-19 up to 300 days. The antibody and cellular responses were tightly interconnected, their magnitude inversely correlated with infection probability. Similar maximal level of protection was reached by individuals positive for both types of responses and by individuals with antibodies alone. Meanwhile, T cells in the absence of antibodies provided an intermediate level of protection. The real-world data on the protective effects of T cells have important implications for T cell immunology and development of the strategies to fight the pandemic.

## Introduction

Severe acute respiratory syndrome coronavirus 2 (SARS-CoV-2), a novel member of the *Betacoronavirus* genus, was identified as a causative agent of a new coronavirus disease 2019 (COVID-19) in December 2019 [1]. Since that, COVID-19 became a worldwide pandemic accounting for over 230 millions of detected cases with a total number of deaths exceeding four million. Significant fractions of the populations in many countries have been infected and cleared of the virus [2, 3]. Exposure of individuals to the virus or its individual antigens (in the case of vaccination) triggers an adaptive immune response including the development of virus-specific effector and memory T cells and the production of antibodies specific to the epitopes of SARS-CoV-2 proteins. It has been shown that immunoglobulins G (IgGs) recognizing the coronavirus spike (S) protein possess virus-neutralizing ability, suggesting that these antibodies could protect from reinfection [4–6].

In addition, growing evidence convincingly demonstrates the important role of T cells in COVID-19. Severe COVID-19 cases are characterized by lymphopenia, affecting both CD4^+^ and CD8^+^ T cells [7]. SARS-CoV-2–specific T cells generated in convalescent patients can contribute to rapid viral clearance and protection from the reinfection [7–10]. Moreover, recent studies revealed the persistence of SARS-CoV-2-specific CD4^+^ and CD8^+^ T cells up to 10 months after infection [11, 12]. Interestingly, CD4^+^ T cells specific to SARS-CoV-2 were also detected in unexposed healthy individuals [8, 13]. These cross-reactive T cells represent memory cells formed after the exposure to other members of the coronavirus family [14]. It has been shown that the presence of pre-existing cross-reactive CD4^+^ T cells accelerates the immune response in SARS-CoV-2 infection and BNT162b2 vaccination [15]. Using rhesus macaques model it was shown that the depletion of T cells (CD4^+^, CD8^+^ or both) led to impaired recovery from acute SARS-CoV-2 infection, while it did not affect the probability of infection [16]. Recently, a prospective study in the UK cohort of emergency workers demonstrated that high levels of SARS-CoV2-specific T cells were correlated with the increased protection from COVID-19, including in participants with low levels of antibodies, but not in the total absence of humoral response [17].

However, the levels of the antibody and T cell responses vary considerably from person to person and may substantially decrease over time [18]. These facts raise an important question: what levels of T cell response and IgG titers are sufficient to protect a person from the infection? The definitive answer to this question can be obtained at the population level from measurements of the immune responses in a large group of individuals followed by tracing of the infection rates in this group.

To find a link between the levels of anti-SARS-CoV-2 antibody response/T cell immune response and protection against the infection, we conducted a prospective study based on the evaluation of the virus-specific Ig levels and virus-specific T cell response in a cohort of 5 340 Moscow residents. Specifically, we evaluated the anti-SARS-CoV-2 IgM/IgG titers and the frequencies of the T cells specific to nucleocapsid (N), membrane (M), and S proteins of SARS-CoV-2, using interferon γ (IFNγ) ELISpot. Furthermore, we assessed the fractions of the virus-specific CD4^+^ and CD8^+^ T cells using intracellular staining of IFNγ and interleukin 2 (IL2) followed by flow cytometry. Finally, we monitored the participants for up to 300 days and analyzed the post-inclusion COVID-19 rates as a function of the assessed antibody and T cell response levels.

Here, we report that both antibody and T cell levels correlated with protection from the SARS-CoV-2 infection. Individuals with strong antibody and cellular responses had very small risk of infection. Surprisingly some level of antibody-mediated protection was evident even at the low titers of antibodies which are considered to be below the cut-off in the standard clinical tests. Overall, the magnitudes of SARS-CoV-2–specific T cell and antibody responses were strongly correlated. However, in a number of individuals in our cohort only one of these two types of response could be detected allowing us to separately analyze the impact of two branches of immunity on protection from COVID-19. The presence of the antibody-only response provided high protection on par with that provided by the simultaneous involvement of both types of response. In contrast, T cells alone provided only intermediate levels of protection from the infection though rates of infection were significantly different from that of the non-immune group. These results provide real world data on the roles of two branches of immunity in disease control and have important implications for public health policy concerning development of recommendations for clinical testing and re-vaccination of individuals who were earlier vaccinated or recovered from COVID-19.

## Materials and methods

### Ethics

This study was approved by the Moscow City Ethics Committee of the Research Institute of the Organization of Health and Healthcare Management and performed according to the Helsinki Declaration. All participants provided their written informed consent. The study was registered on ClinicalTrials.gov (Identifier: NCT04898140).

### Participant inclusion

The study cohort consisted of individuals above 18 years old who had voluntarily come to one of four Moscow City Clinics for routine testing for COVID-19 antibodies and agreed to participate in the study. No specific inclusion or exclusion criteria beyond age were applied. After providing the written informed consent, the individuals hand-filled a questionnaire containing information about their demographics, health, marital and social status, and self-estimated previous COVID-19 status or possible contacts with COVID-19 positive individuals.

### Study subjects observation

Information about individuals’ medical histories in relation to COVID-19 pandemics was obtained from the Moscow State COVID-19 registry, which contains information about all individuals who have been tested positive for COVID-19 by means of either PCR or serology tests, persons who participated in one of the COVID-19 vaccine clinical studies, or who were vaccinated against COVID-19 with an approved vaccine. This registry also contains information about the dates when the patients were diagnosed for COVID-19, the types of treatment they received, and the outcomes of the disease. This information was used both for analysis of the data from the individuals who were SARS-CoV-2–positive prior to inclusion in the study and for identification of the individuals who were infected post-inclusion.

### Blood collection and PBMC isolation

Peripheral blood was collected into two 8-mL VACUTAINER® CPT™ tubes with sodium citrate (BD, USA) and was processed within two hours after venipuncture. Peripheral blood mononuclear cells (PBMC) were isolated according to the manufacturer’s standard protocol by centrifugation at 1 800–2 000g for 20 min with slow brake at room temperature (RT). After centrifugation, an aliquot of blood plasma was collected into a 1.5-mL microcentrifuge tube and stored at –20 °C or lower, while PBMC were collected into a 15-mL conical tube, washed twice with phosphate buffered saline (PBS, PanEco, Russia) with EDTA at 2 mM (PanEco, Russia), counted, and used for IFNγ ELISpot assay and flow cytometry with intracellular cytokine immunostaining. PBMC with a viability level ≥70% were taken into the study. For serum isolation, peripheral blood was collected into S-Monovette 7.5-mL Z tubes (Sarstedt, Germany).

### SARS-CoV-2–specific antibodies and virus-neutralizing activity of plasma

We evaluated the titers of the virus-specific IgM and IgG antibodies in blood serum using an automated CL-series chemiluminescent immunoassay analyzer with compatible reagent kits (Mindray, China); here and below ‘IgG’ and ‘IgM’ refer to results obtained with this method, if not mentioned otherwise. For a separate representative group of participants, we measured antibodies specific to SARS-CoV-2 spike (S) and nucleocapsid (N) proteins using the SARS-CoV-2-IgG-EIA-BEST ELISA kit (Vector-Best, Russia) and the automated ARCHITECT i1000SR analyzer with compatible reagent kit (Abbott, USA), respectively, according to the manufacturer’s standard protocol; additionally, we measured the virus-neutralizing activity of plasma by microneutralization assay using SARS-CoV-2 (hCoV-19/Russia/Moscow_PMVL-1/2020) in a 96-well plate and a 50% tissue culture infective dose (TCID50) of 100 as described in [6] with plasma dilutions of 10, 20, 40, 80, 160, 320, 640, and 1 280 times.

### Flow cytometry

PBMC were plated into 96-well U-bottom plates at a concentration of 1×10^6^ cells per well in 100 µL of serum-free AIM-V medium (ThermoFisher Scientific; USA) supplemented with 1X AlbuMAX (ThermoFisher Scientific; USA), 2 mM L-glutamine, 50 μg/mL streptomycin, and 10 μg/mL gentamicin. The cells were stimulated with a mixture of SARS-CoV-2 PepTivator S, S1, S+, N, and M peptide pools (each 1 μg/mL, Miltenyi Biotec, Germany) for 3 h; then brefeldin A (BrA, Merck, Germany) was added to a final concentration 10 μg/mL. An equal amount of BrA was added to non-stimulated negative control cells, as well as to positive control cells stimulated with ionomycin at 1 µM (Merck, Germany) and phorbol-12-myristate-13-acetate (Merck, Germany) at 40 nM for 2 h. After the BrA addition, the plates were incubated for 14–16 h at 37 °C in 5% CO2 atmosphere, and then the cells were washed with PBS, blocked with 5% normal mouse serum (NMS, Capricorn Scientific, Germany), and stained with anti-CD45-PerCP (clone HI30, BioLegend, USA), anti-CD3-APC (clone OKT3, BioLegend, USA), anti-CD4-FITC (clone OKT4, BioLegend, USA), and anti-CD8a-PE (clone HIT8a, BioLegend, USA) conjugates for 15 min, washed with PBS, and fixed with 2% paraformaldehyde (Merck, Germany) at 4 °C for 20 min. After fixation, the cells were washed with 0.2% saponin in PBS (Merck, Germany), blocked with 5% NMS, and stained with anti-IFNγ-PE/Cy7 (clone 4S.B3, BioLegend; USA) and anti-IL2-APC/Cy7 (clone MQ1-17H12, BioLegend; USA) conjugates for 40 min in 0.2% saponin in PBS. We analyzed stained cells using FACSCanto, FACSCanto II, FACSCAria SORP (BD Biosciences, USA), and Novocyte 2060 (ACEA Biosciences, Agilent Technologies, USA) instruments equipped with 488-nm and 640-nm lasers with suitable sets of optical filters. We analyzed data using FlowJo software (BD Biosciences, USA). For each specimen at least 100 000 single CD3^+^CD45^+^ events were recorded. The compensation matrix was calculated automatically by the FlowJo software using single-stained CompBeads (BD Biosciences, USA).

### IFNγ ELISpot assay

We performed an IFNγ ELISpot assay using the Human IFNγ Single-Color ELISPOT kit (CTL; USA) with a 96-well nitrocellulose plate pre-coated with human IFNγ capture antibody according to the manufacturer’s protocol. Briefly, 3×10^5^ freshly isolated PBMC in serum-free CTL-test medium (CTL, USA), supplemented with Glutamax (ThermoFisher Scientific, USA) and penicillin/streptomycin (ThermoFisher Scientific, USA), were plated per well and incubated with SARS-CoV-2 PepTivator N or M or a mixture of S, S1, and S+ peptide pools (Miltenyi Biotec, Germany) at a final concentration of 1 μg/mL each at a final volume of 150 µL/well. Additionally, cells were incubated with media only (negative control) or phytohaemagglutinin (Paneco, Russia) at a final concentration of 10 µg/mL (positive control). Plates were incubated for 16–18 h at 37 °C in 5% CO2 atmosphere. The plates were washed twice with PBS, then washed twice with PBS containing 0.05% Tween-20, and incubated with biotinylated anti-human IFNγ detection antibody for 2 h at RT. Plates were washed three times with PBS containing 0.05% Tween-20 followed by incubation with streptavidin-AP for 30 min at RT. Spots were visualized by incubation with the substrate solution for 15 min at RT. The reaction was stopped by a gentle rinse with tap water. We air-dried plates overnight at RT and then counted spots using an automated spot counter CTL ImmunoSpot Analyzer and ImmunoSpot software (CTL; USA). Samples in which the negative control was greater than 10 spots and/or the positive control was less than 20 spots were considered as invalid.

### Statistical analysis

Statistical analysis was performed with the Python3 programming language with *numpy*, *scipy*, *pandas* and *lifelines* packages. The Fisher exact test (two-tailed) was used to compare qualitative parameters between independent groups of individuals; the significance level α for *p*-values was set to 0.05. The Mann–Whitney U test (two-sided) was used for comparing distributions of quantitative parameters between independent groups of individuals. To control for type I error, we calculated false discovery rate *q*-values using the Benjamin– Hochberg (BH) procedure and set a threshold of 0.05 to keep the positive false discovery rate below 5%. The Kaplan–Meier estimator technique was used to assess the effects of different types of immunity on the probability of getting ill, with the log-rank test used to compare the survival distributions of different groups (see Supplementary Materials Section 3 for more details). In all figures, for simplicity, *p*-values are ranked by significance levels using the following labels: 1.00e-02 < *p*-values ≤ 5.00e-02 are marked with ‘*’; 1.00e-03 < *p*-value ≤ 1.00e-02 with ‘**’; 1.00e-04 < *p*-value ≤ 1.00e-03 with ‘***’; and *p*-value ≤ 1.00e-04 with ‘****’. All boxplots presented below are standard box-and-whisker plots […].

### Positivity criteria

We set positivity thresholds at 10 AU/mL and 1 Cutoff Index (COI) for IgG and IgM tests, respectively, and performed the tests using a CL-series chemiluminescent immunoassay analyzer (Mindray; China) according to the manufacturer’s standard instructions.

For ELISpot and flow cytometry intracellular immunostaining tests, no pre-defined positivity criteria existed. In order to create positivity criteria which would optimally discriminate between individuals who had been diagnosed with COVID-19 and those who had not (similar to antibody test positivity criteria), we first split the participants into two groups. The first group consisted of individuals with no diagnosed COVID-19 or COVID-19 contacts according to state authorities, with no self-reported COVID-19 or COVID-19 contacts, with no reported ARI symptoms six months prior to the inclusion, and with IgG < 1 AU/mL, IgM < 1 COI. The second group consisted of the individuals with PCR-confirmed COVID-19 who recovered before the study, were registered by the state authority, and self-reported COVID-19. We then developed the criteria which would allow us to reach the optimal separation between these groups (see Supplementary Materials for details).

## Results

### Cohort characteristics

In this prospective non-randomized study, we analyzed the levels of SARS-CoV-2 specific antibodies and the frequencies of the virus-specific T cells in peripheral blood among 5 340 individuals recruited from the Moscow general population. The study design is schematically represented in Figure 1A. All individuals enrolled in the study were Moscow residents over 18 years old, females (63.8%) and males (36.2%), with the mean age being 43±14 and 42±13 years, respectively (Figure 1B). The cohort recruitment lasted for 10 weeks (October to December, 2020). At the moment of inclusion 854 participants (17%) had previously clinically confirmed COVID-19 infection, 81 more (2%) were diagnosed at the time of inclusion. The post-inclusion observation continued until the last week of August, 2021. The COVID-19 status of all individuals enrolled in the study by the end of observation is presented in Figure 1C.

**Figure 1.**
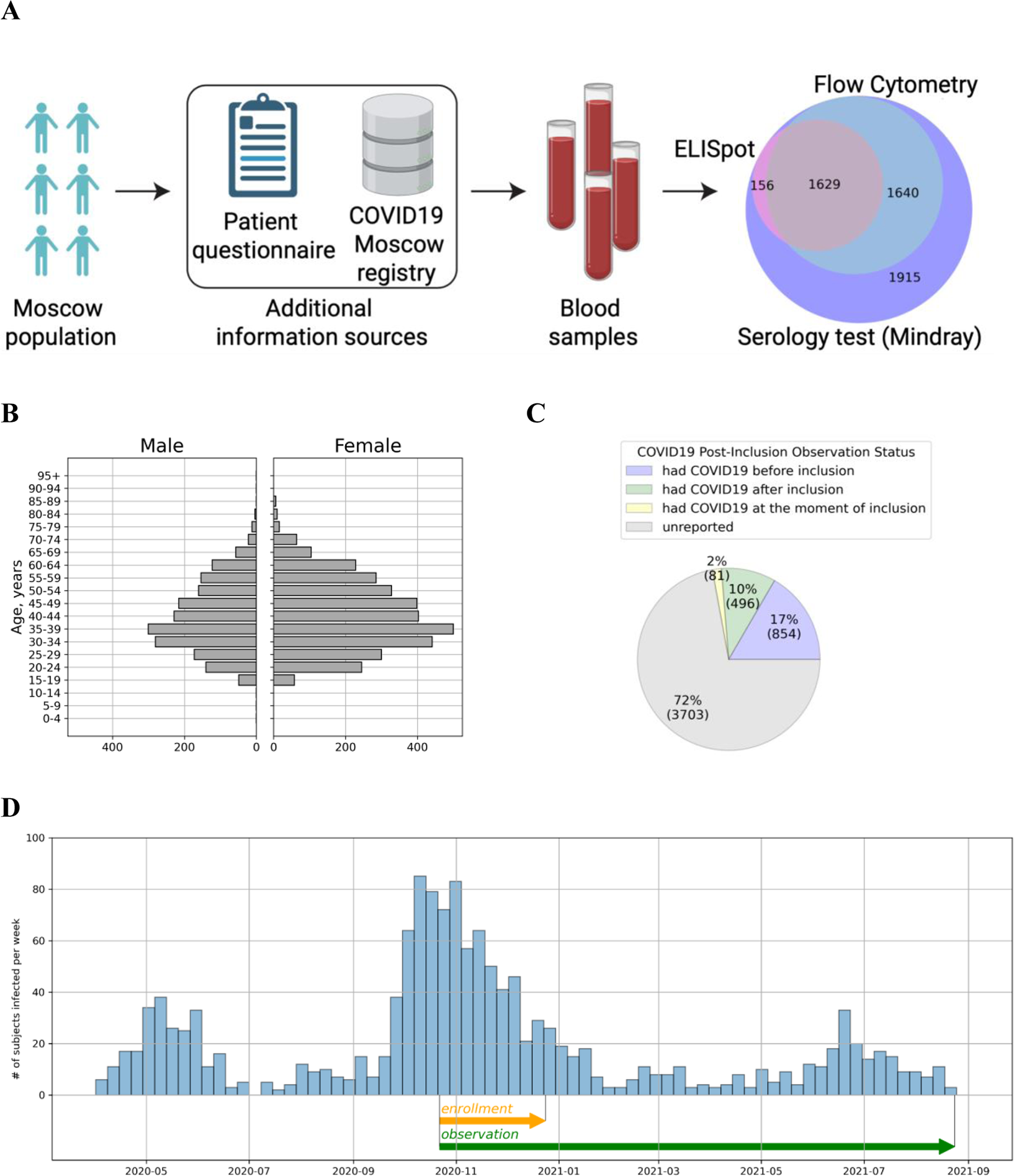
Study overview and experimental cohort description. **(A)** Schematic study design. Volunteers were tested for SARS-CoV-2–specific antibodies and T cell response. T cellular immune response for SARS-CoV-2 proteins was assessed from IFNɣ ELISPOT and flow cytometry. COVID-19 status of the participants was evaluated on the basis of questionnaire responses and the Moscow State COVID-19 registry. **(B)** Age and Sex distribution of volunteers included in the study. **(C)** COVID-19 status of volunteers included in the study according to the Moscow State COVID-19 registry provided by the Moscow Department of Healthcare. **(D)** COVID-19 cases among study participants per week from April 2020 to August 2021 according to the Moscow State COVID-19 registry provided by the Moscow Department of Healthcare. Enrollment and observation periods of the study are shown as orange and green arrows, respectively.

### Antibody response

Antibody levels were measured for 5 328 participants. At the time of inclusion, 1 382 (26%) individuals were positive for SARS-CoV-2-specific immunoglobulins M (IgMs) and 2 455 (46%) were positive for the SARS-CoV-2–specific immunoglobulins G (IgGs) (Figure 2A, B). It is noteworthy that the average proportion of IgG-positive individuals gradually increased from 37% in the first week to 55% in the last week of recruitment (see Supplementary Figure S1 for details). To evaluate the stability of the immune response we selected a subgroup of 854 participants who had clinically confirmed COVID-19 (by PCR) at various time points prior to inclusion in the study. We found that IgM titers in the blood serum of these recovered individuals considerably decreased after the 60th day post-disease-onset (Figure 2C). At the same time, IgG titers stayed relatively high and largely unaltered up to 270 days after the disease (Figure 2D) — the longest time between disease confirmation and blood collection in our study. Within a smaller representative group of individuals of the cohort (n = 180) we evaluated virus-neutralizing activity (VNA) of peripheral blood plasma, as well as titers of S- and N-protein–specific IgG antibodies. We detected a strong correlation between IgG titers measured by the assay used in this study, on the one hand, and plasma VNA, as well as both S- and N-protein–specific antibodies, on the other (see Supplementary Figure S2 and Supplementary Materials for details).

**Figure 2.**
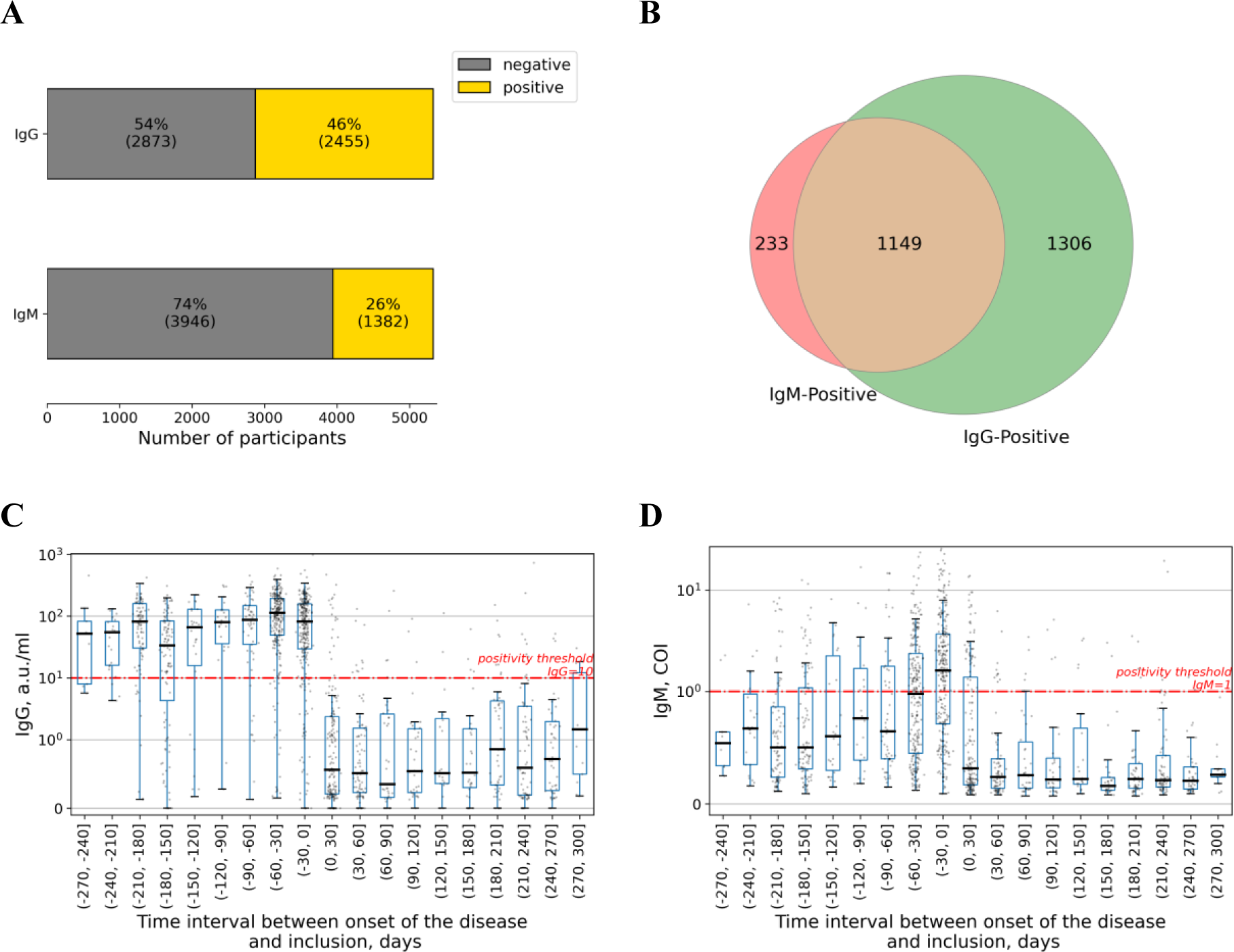
Evaluation of COVID-19-specific antibody immunity. **(A)** Percentages of patients exceeding a positivity threshold for IgM and IgG. **(B)** Venn diagram showing relation in positivity between IgG and IgM antibody types. **(C, D)** Time dependence of the IgG and IgM levels are shown in **(C)** and **(D),** respectively. Each dot represents a single patient. Time is counted from the date of disease onset according to the Moscow State COVID-19 registry to the day of inclusion in the study. Time interval presented in each boxplot is 30 days. Red line represents a positivity threshold. COI, Cutoff Index.

### T cell response

We analyzed the frequencies of the T cells specific to nucleocapsid (N), membrane (M), and spike (S) proteins of SARS-CoV-2 in peripheral blood using ELISpot for 1 785 participants, as well as the frequencies of virus-specific CD4^+^ and CD8^+^ T cells using flow cytometry for 3 269 participants, with analyses using both methods performed in 1 629 participants (Figure 1A). For this purpose, we used peptide pools (15-mer peptides overlapping by 10 residues and spanning the SARS-CoV-2 M, N, and S proteins) and a stimulation protocol described elsewhere [8,19,20]. Both ELISpot and flow cytometry showed that approximately half of analyzed individuals demonstrated the presence of the T cell response against SARS-CoV-2 antigens that was consistent with the level of the antibody response in the cohort (Figure 3A, B). Overall, 1 145 (64.1%) individuals had SARS-CoV-2–specific T cells with response to at least one of the SARS-CoV-2 proteins (N, M, or S) as measured with ELISpot, including 692 (38.8%) with immune responses to all three proteins (Figure 3C). Meanwhile, as analyzed with intracellular immunostaining and flow cytometry, 2,217 (67.8%) participants had SARS-CoV-2–specific CD4^+^ T cells expressing either interleukin 2 (IL2) only, interferon γ (IFNγ) only, or both cytokines simultaneously, with 1 095 (33.5%) participants having all three cell populations (Figure 3D). All the metrics of T cell immunity appeared to be relatively stable up to 270 days post COVID-19 (Figure 3E,F; Supplementary Figure S3). Meanwhile, CD8^+^ T cells activated upon stimulation, as expected, were represented predominantly by the cell population expressing only IFNγ (see Supplementary Figure S4H, see Supplementary Materials for details).

**Figure 3.**
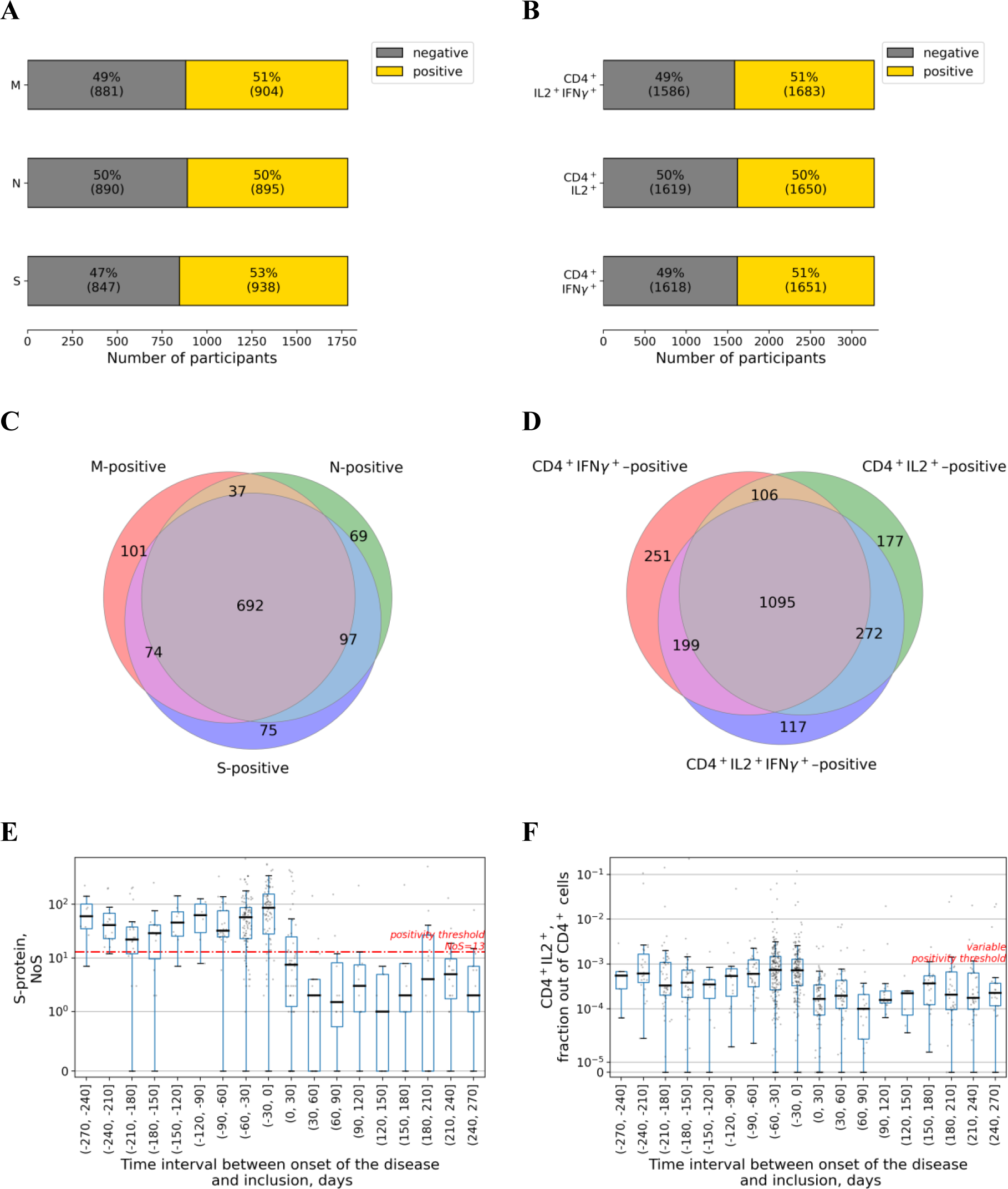
Evaluation of COVID-19-specific T cell immunity. **(A)** Percentages of patients exceeding a positivity threshold for M, N, S in the ELISpot assay. **(B)** Percentages of patients exceeding a positivity threshold for percentage of cells expressing both IL2 and IFNγ, or either of these cytokines in the flow cytometry assay. **(C)** Venn diagram showing relation in positivity between different SARS-CoV-2 proteins in the ELISpot assay. **(D)** Venn diagram showing relation in positivity between expression of different cytokines in response to activation with SARS-CoV-2 proteins in the flow cytometry assay. **(E)** Time dependence of the spots numbers for S-protein in the ELISpot assay. Each dot represents a single patient. Time is counted from the date of disease onset according to the Moscow State COVID-19 registry to the day of inclusion in the study and thus serology testing. Time interval presented in each boxplot is 30 days. Red line represents a positivity threshold. NoS, number of spots. **(F)** Time dependence of the fraction of CD4^+^ T cells expressing IL2 out of total CD4^+^ cells in the flow cytometry assay. Each dot represents a single patient. Time is counted from the date of disease onset according to the Moscow State COVID-19 registry to the day of inclusion in the study and thus of serology testing. Time interval presented in each boxplot is 30 days. The positivity threshold was variable (see Supplementary Materials for more details) and thus not given here.

### Correlation between different metrics of immune response

The frequencies of SARS-CoV-2 –specific T cells measured by ELISpot were compared with the frequencies of activated T cell subpopulations ascertained from intracellular immunostaining with flow cytometry. As expected, a strong correlation of the data obtained by different methods was observed (SupplementaryFigure S4; see Supplementary Materials for details). Additionally, we found a strong correlation between IgG titers and T cell frequencies as determined by both ELISpot and flow cytometry (Figure 4; see Supplementary Materials for more details). It is worth noting that this correlation was found in the cases of N-, M-, and S-protein–specific T cells, as well as for different populations of CD4^+^ T cells, but not in the case of CD8^+^ T cells (SupplementaryFigure S4).

**Figure 4.**
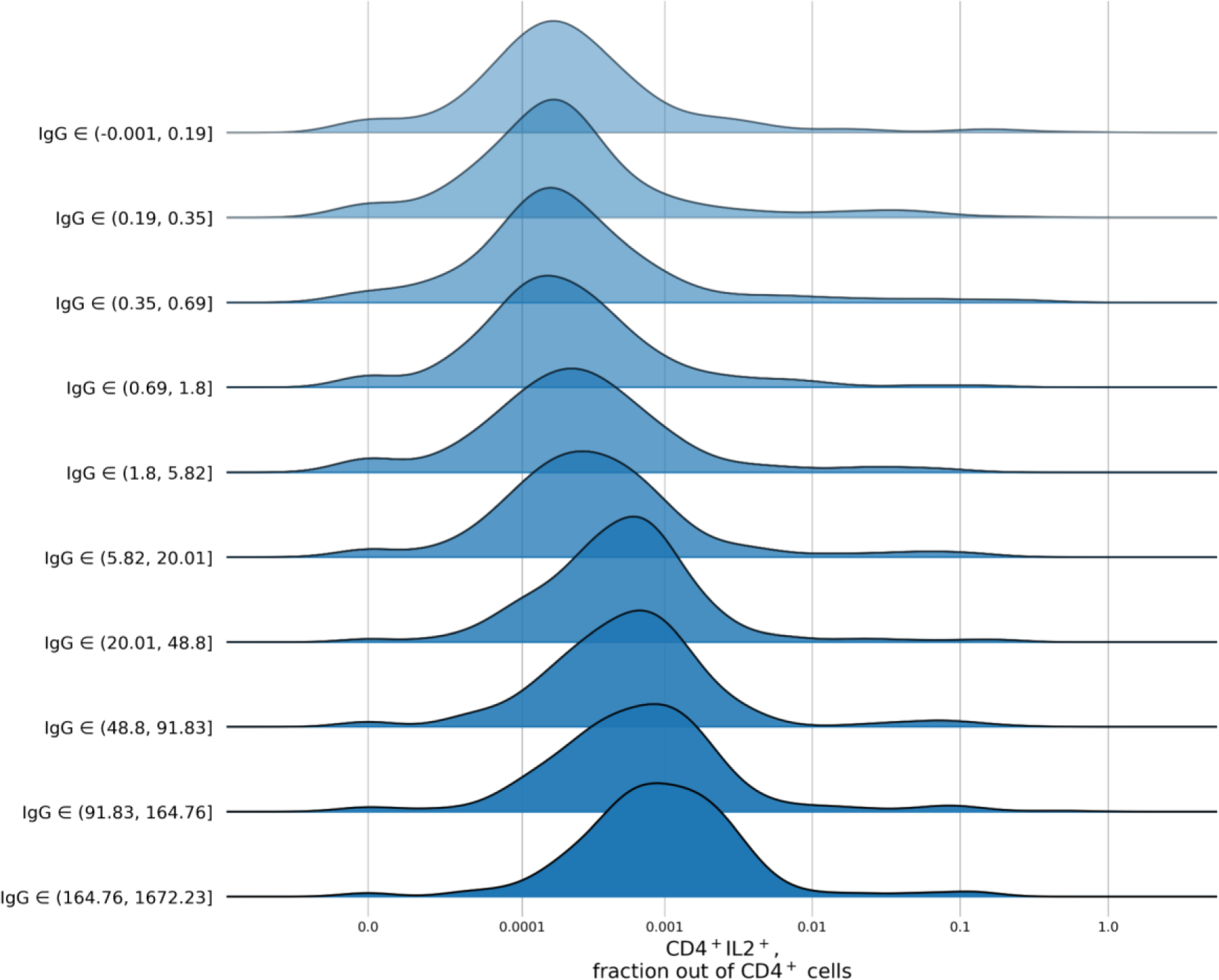
Ridgeline plots for T cell immunity metrics as a function of antibody levels. Distribution per decile of IgG of flow cytometry CD4^+^ cells expressing the IL2 fraction out of the total number of CD4^+^ cells. For patients from each IgG decile, the plot shows the kernel-density estimate for the T cell fraction, using the Gaussian kernel

### Post-inclusion observation

For the cohort of participants, we analyzed the post-inclusion COVID-19 rates. To evaluate the effects of different levels of IgG antibodies or T cell responses on the susceptibility to SARS-CoV-2 infection among participants, we employed the Kaplan-Meyer estimator method. Among the 4,027 participants who were eligible for the post-inclusion observation (see Supplementary Materials for more details on Kaplan Meyer curves analysis), a total of 496 post-inclusion cases of COVID-19 were registered. For each of the studied parameters of immune response, all participants were divided by the quantiles depending on the levels of their responses, and corresponding Kaplan-Meier curves for each group were generated and analyzed (see Supplementary Materials for more details on Kaplan Meyer curves analysis).

In case of virus-specific IgG levels (Figure 5A), individuals with IgG titers under 0.29 AU/mL, representing the Q1 quantile, were characterized by the highest infection chances (approximately 22%) by the end of the observational period. Infection chances for individuals in the Q2 and Q3 quantiles (with IgG titers ranging 0.29–0.97 and 0.97–8.33 AU/mL, respectively) were slightly lower (approximately 16.5%), compared with Q1, and the difference was statistically significant (log-rank *p*-value < 0.05). Finally, individuals with IgG titers higher than 8.32 AU/mL demonstrated considerably lower infection chances - approximately 6% and 2.5% for the Q4 (8.33–66.76 AU/mL) and Q5 (>66.76 AU/mL) quantiles, respectively. The 4th and 5th IgG quantiles which demonstrated the highest degree of protection also had the highest levels of plasma virus-neutralizing activity (Supplementary Figure S2B). Meanwhile, Q3 quantile with the intermediate infection chances had significant increase of virus-neutralizing activity over Q1. However, there was no difference in VNA between the Q1 and Q2 quantiles.

**Figure 5.**
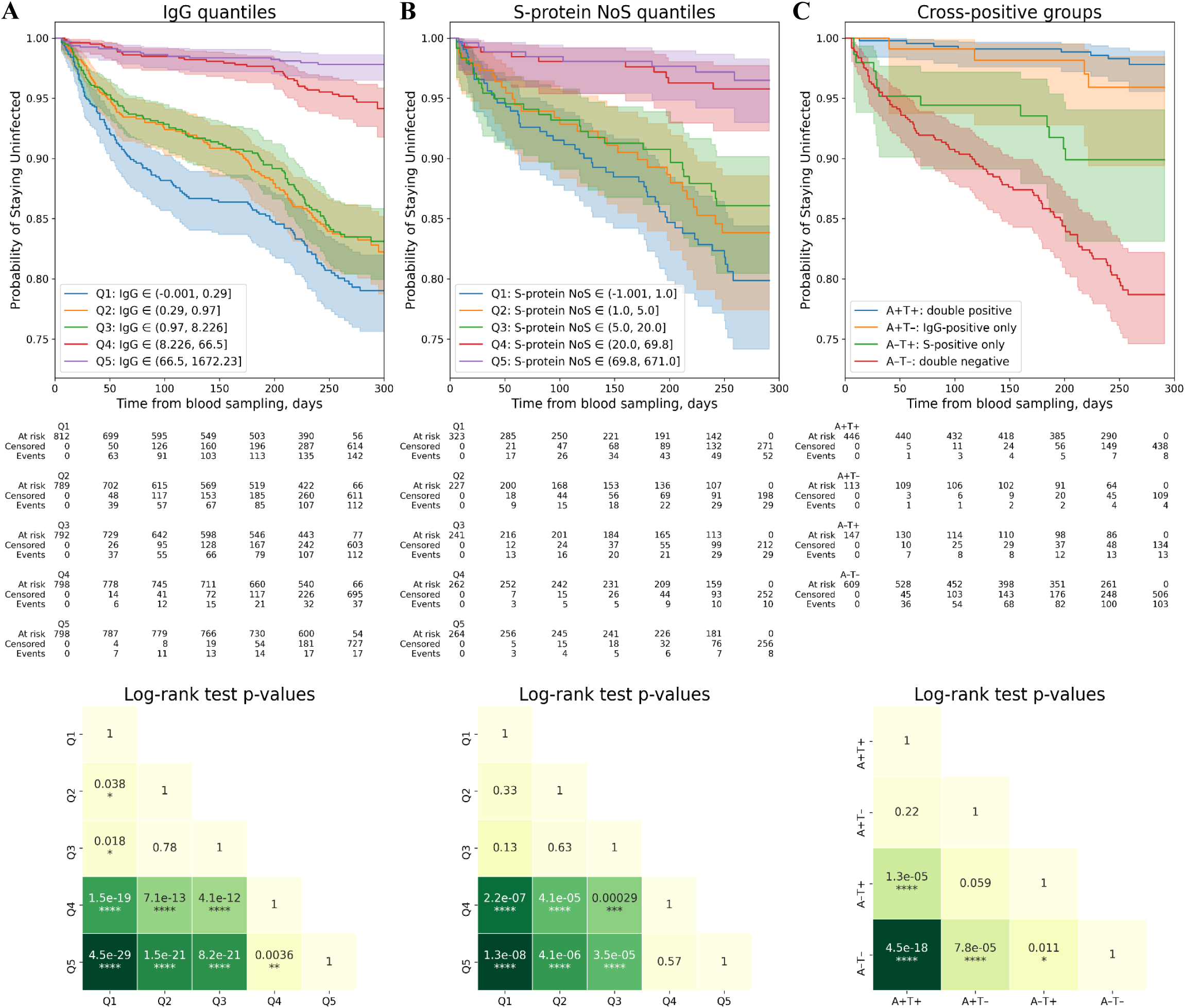
Evaluation of effects of antibody immunity and T cell immunity on COVID-19 infection rates. **(A) Top**: Kaplan-Meyer curves for patients with different IgG levels. The patients were split into five nearly equal groups by quantiles of IgG from Q1 to Q5, and a Kaplan-Meyer curve was built for each group (see Supplementary Materials for more details on Kaplan Meyer curves analysis). **Bottom**: log-rank test *p*-values for pairwise comparison of all five groups selected by quantiles. **(B) Top**: Kaplan-Meyer curves for patients with different number of spots (NoS) for S-protein specific T cells estimated using ELISpot. The patients were split into five nearly equal groups by quantiles of cell fraction from Q1 to Q5, and a Kaplan-Meyer curve was built for each group (see Supplementary Materials for more details on Kaplan Meyer curves analysis). **Bottom**: log-rank test *p*-values for pairwise comparison of all five groups selected by quantiles. **(C) Top**: Kaplan-Meyer curves for patients with different positivity by IgG levels and NoS for S-protein specific T cells. The participants were split into four groups: positive only by antibodies (A+T-), positive only by S-protein specific T cells (A-T+), double-positive (A+T+), and double-negative (A-T-), with the positivity criteria discussed above used for the evaluation, and a Kaplan-Meyer curve was built for each group (see Supplementary Materials for more details on Kaplan Meyer curves analysis). **Bottom**: log-rank test *p*-values for pairwise comparison of all four groups selected by positivity.

An almost binary relationship was observed between infection chances and the frequencies of virus–specific T cells identified by ELISpot (Figure 5B and Supplementary Figure S5). For all the SARS-CoV-2 proteins analyzed (i.e., M, N and S), individuals comprising Q4 and Q5 quantiles were characterized by the highest protection against the infection, while the remaining quantiles (Q1, Q2 and Q3) were similar and demonstrated no considerable protection. The results obtained demonstrated that the maximal protection was observed if the number of spots was above 15, 7 and 20 for M, N, and S-protein specific T cells, respectively.

In contrast to the ELISpot data, the results of the T cell response by flow cytometry demonstrated a gradual relationship between the parameter’s value and infection chances, with the infection chances getting lower gradually as the values getting higher. This was the case for the frequencies of the CD4^+^ T cells expressing IFNγ, IL2, as well as both cytokines simultaneously (Supplementary Figure S5). The same tendency was observed for the CD8^+^ T cells expressing IFNγ; however, these data were less statistically significant (Supplementary Figure S5). Nevertheless, if three CD4^+^ T cell subpopulations (namely, expressing IFNγ, IL2, or both cytokines) were combined, the relationship with infection chances transformed into binary, with the protectivity against the infection being the highest among individuals comprising Q3, Q4, and Q5 quantiles. Thus, the maximal protection is achieved when the frequency of virus-specific CD4+ T cells is higher than 67 cells per 100 000 total peripheral blood CD4+ T cells (Supplementary Figure S5).

The same method of group comparison, using Kaplan-Meyer curves, was employed to separate the effects of cellular and antibody responses on protection against SARS-CoV-2 infection. For this purpose, the participants were split into four groups: positive only by antibody response (A+T-), positive only by some metric of T cell immune response (A-T+), double-positive (A+T+), and double-negative (A-T-), with the previously estimated positivity criteria used for the evaluation (see Materials and Methods section). Such analysis was performed for all metrics of T cell response used in this study, except for CD8^+^ T cells since it was impossible to develop a reliable positivity criterion (Figure 5C and Supplementary Figure S6). For all of the metrics, the A-T-group had the highest infection chances, while the strongest protection was observed in the A+T+ group and in the A+T-group, the latter two groups being statistically indistinguishable (Figure 5C, Supplementary Figure S6). For all studied metrics of T cell immune response, A-T+ group demonstrated intermediate protection that was significantly lower than both the A+T+ and the A+T-groups but for some metrics it was significantly higher than in the A-T-group (Figure 5C, Supplementary Figure S6). In particular, the protection provided by the T cells in the absence of antibodies was observed when the response was estimated by the numbers of N- and S-protein specific T cells by ELISpot (Figure 5C, Supplementary Figure S6A, B). The tendency for the increased protection was observed for the frequencies of the CD4^+^ T cells expressing IFNγ, IL2, or both cytokines (Supplementary Figure S6D, E, F), but the statistical significance was achieved only if these subpopulations were combined (Supplementary Figure S6G). However, it should be emphasized that individuals single-positive for N- and S-protein specific T cells, as well as for combined virus-specific CD4^+^ T cells, were characterized by higher IgG levels than individuals of the A-T- group, although the antibody levels were below the positivity cut-off value of 10 AU/mL set by the serology test manufacturer (Supplementary Figure S7).

## Discussion

With the progression of the COVID-19 epidemic, a growing number of individuals developed immune responses against SARS-CoV-2. Prospective studies in humans [21–23] and studies using primate models with SARS-CoV-2 re-challenge [24–26] demonstrated that acquired immune response after COVID-19 provides protection from the infection. However, the strength of the immune response in humans is very diverse. Therefore, it remains to be understood what immune response metrics correlate with the protection. To answer this question, in the present study we investigated the protective potential of the antibody and T cell immune responses against the SARS-CoV-2 in the context of the COVID-19 epidemic in Moscow between October 2020 and August 2021.

A constant increase in the average proportion of IgG-positive individuals which we observed during the recruitment phase of the study is most likely explained by the growing number of COVID-19 cases during the second wave in Moscow, coinciding with the time of the cohort recruitment [https://www.who.int/countries/rus/].

By measuring antibody and T cell responses in participants who had PCR-confirmed COVID-19 at different time points before enrollment we were able to estimate the longevity of immune response. We showed that IgM titers rapidly decreased after two months post-infection, whereas both IgG and virus-specific T cells could be found in the peripheral blood up to 270 days post-infection with no apparent decrease in median response rate. These data are in agreement with the results of studies analyzing the immune response dynamics in patients recovered after COVID-19 [27–29].

Here, we report a strong correlation between frequencies of SARS-CoV-2–specific T cells evaluated with ELISpot and those evaluated with flow cytometry. These results are not surprising, considering that both methods detect cytokine expression in the activated T cells upon their stimulation with the specific antigens [30]. Importantly, IgG titers are highly correlated with the frequencies of SARS-CoV-2–specific T cells, indicating, as expected, that antibody and cellular responses are closely interconnected and are induced concurrently in the vast majority of individuals. This correlation was found even at the IgG values below the seropositivity cut-off.

By analyzing the post-inclusion COVID-19 rates we were able to estimate which metrics of the immune response correlated with the protection. We found that IgG titers and all studied parameters of the T cell response estimated in the study negatively correlated with the SARS-CoV-2 infection probabilities. T cell response as measured by ELISpot was generally characterized by the binary relationship between the response level and infection probabilities. This means that the protection against the SARS-CoV-2 infection was the same for all individuals who demonstrate the frequency of the SARS-CoV-2 specific T cells in peripheral blood surpassing a particular threshold. However, a different type of relationship between protectivity against the infection and parameter level was observed in the case of IgG titers. We identified three groups of individuals characterized by different infection chances. Individuals with IgG titer under 0.29 AU/mL were characterized by the highest infection chances, while the lowest infection chances were associated with values above 8.33 AU/mL, regardless of the exact value. At the same time, infection chances for individuals with IgG titers in the range 0.29–8.33 AU/mL were slightly lower than those for the group with the lowest IgG values, but at the same were considerably higher than in the group with the highest IgG titers. Moreover, we found significant virus-neutralizing activity among individuals with IgG titers in the range 0.97–8.33 AU/mL which is below the seropositivity cut-off of 10 AU/mL set by the manufacturer. This finding may have several possible explanations. This group with intermediate values of IgG may be represented by individuals recovered after asymptomatic or mild COVID-19, who have been shown to develop no antibody response or a rapidly decreasing one [31, 32]. Alternatively, these individuals may have IgG antibodies that developed as a result of a previous infection with the “common cold” coronaviruses and that are cross-reactive to SARS-CoV-2 [8].

To separate the effects of T cell and antibody responses on protection against SARS-CoV-2 infection we split the participants into four groups: positive by antibodies only, positive by some metric of T cell immune response, but negative for antibodies, double-positive, and double-negative. Two groups were characterized by the highest protection levels: individuals positive for both types of responses and individuals with antibody response devoid of T cells

It is noteworthy that, despite the fact that the COVID-19 rates in these groups were very small - approximately 1%, the rare infection cases were still detected. At the same time, individuals with T cell response but absence of antibodies demonstrated intermediate protection levels which nevertheless were significantly different from the individuals without either type of immunity. Not all T cell response metrics analyzed in this study were found to correlate with the protection against the infection. Statistically significant protection compared to double-negative individuals was observed for N- and S-protein specific T cell response (measured by ELISpot), and for overall SARS-CoV-2 specific CD4^+^ T cell response (measured with flow cytometry). This modest effect of T cells on protection from the infection possibly originates from the need for T cells to undergo proliferation for several days and migrate from peripheral blood to the site of infection which leads to an unavoidable delay in the viral clearance. In contrast, antibodies have instantaneous activity and, thus, provide for the faster response. Still, it should be noted that individuals single-positive for any of the above mentioned T cell response metrics were characterized by higher IgG titers than individuals without either type of immunity, although the titers were below the positivity cut-off value of 10 AU/mL set by the test manufacturer.

These results are not surprising, considering the strong relationship and proportionality between antibody and T cell immunities found in our study. However, these data, taken together with the high protectivity of the antibody-response-only-positive group, indicate that antibodies better correlate with the protectivity against the SARS-CoV-2 infection. In turn, the results of our study show that IgG evaluation in serum is a more precise method for the prognosis of SARS-CoV-2 infection chances than the methods based on the detection of virus-specific T cells.

The most important role of T cells might be not in protection from the primary infection but in contribution to the viral clearance and thus in managing the disease severity. It was reported that impaired CD8^+^ T cell responses contributed to poor prognosis [33]. On the contrary, a higher proportion of SARS-CoV-2 specific CD8^+^ T cells was observed in patients with milder disease [34]. Moreover, in patients with severe disease T cells were characterized by the exhaustion phenotype and decreased production of IFNγ [35–37]. In addition, COVID-19 patients over 80 years old, who are known to develop severe disease, had decreased CD8+ T cell responses [38].

Our study has several limitations: (i) the cohort analyzed is likely to be non-representative and includes only individuals who have visited outpatient clinics to voluntarily undertake tests for COVID-19 antibodies levels and who agreed to participate in the study; (ii) only a limited fraction of cases were reported to the Moscow State COVID-19 registry, so some COVID-19 cases in our cohort after inclusion remained unanalyzed, though we don’t expect any nonrandom distribution of unreported cases between different groups; and (iii) here we evaluated contributions of systemic antibody and T cell responses to the infectivity probabilities, whereas their local concentrations in respiratory system, where the critical events of the SARS-CoV-2 infection occur, may be different.

Taken together, our data suggest that the contribution of the virus-specific antibody response to protection against the SARS-CoV-2 infection is more pronounced than that of the T cell response. Thus, in order to predict COVID-19 rates on a population level serological testing is advantageous. The data on the specific IgG titers may be instructive for making decisions in personalized healthcare, as well as for the development of public anti-COVID-19 policies.

## Data Availability

Data available within the article and supplementary materials

## Acknowledgements

The authors would like to thank Dr. Leonid Margolis (Eunice Kennedy Shriver National Institute of Child Health and Human Development, National Institutes of Health, Bethesda, MD, USA) and Prof. Michael M. Lederman (Case Western Reserve University/University Hospitals Cleveland Medical Center, Cleveland, Ohio, USA) for their valuable comments and suggestions during experimental design, discussion of the results and manuscript preparation, and Dr. Barry Alpher for assistance in editing and improving the English style of the manuscript. The authors also thank the Moscow Department of Healthcare for the help in organization of the study.

## Author Contributions

G.A.E., I.A.K., A.A.K., M.R.K., D.Y.L., N.B.N., Y.P.R., A.V.S, I.A.V., P.V., and E.V. designed and coordinated the study. I.A.M. performed the statistical analysis and together with E.K., A.N.M., O.M., O.E.M., A.E.P., M.V.P., and I.O.P. performed initial data interpretation. A.A., W.A., A.S.B., A.S.D., I.V.D., I.N.F., A.N.G., O.I.I., A.K., V.V.K., A.K., N.I.K., D.A.L., Y.A.L., A.V.M., E.V.M., A.M., V.V.M., N.E.M., A.N., M.F.N., L.A.O., N.V.P., D.M.P., E.V.R., A.A.S, N.S., A.G.S., Y.S., N.T.S., N.I.S., S.A.S., A.F.S., L.S., A.T., A.V.T., V.M.U., A.S.V., D.A.V., and K.V.Z. performed the experiments. Every author contributed to the initial draft of the manuscript and agreed on submission for publication. All authors interpreted the data, reviewed the manuscript and approved the final version.

## Supplementary Materials

### Supplementary Section 1: Positivity Criteria

For ELISPOT and flow cytometry tests, no pre-defined positivity criteria existed. In order to create positivity criteria which would optimally discriminate between people who were diagnosed with COVID-19 and those who had not been so diagnosed (in a manner similar to antibody test positivity criteria), we first selected two groups of patients. The first group, or ‘negative group,’ consisted of patients with no earlier COVID-19 or COVID-19 contacts reported by state authorities, no self-reported COVID-19 or COVID-19 contacts, no ARI symptoms six months prior to the inclusion reported, with IgG, Mindray < 1 AU/mL, and with IgM, Mindray < 1 COI. The second group, or ‘positive group’, consisted of the patients with PCR-confirmed COVID-19 who recovered before the study, as reported by state authority, and self-reported COVID-19.

There were a total of 401 patients in the negative group, out of whom 209 had ELISpot results and 292 had flow cytometry results, and there were 563 patients in the positive group, out of whom 303 had ELISpot results and 318 had flow cytometry results.

We then developed the criteria which would allow us to reach the optimal separation between two such groups using results of ELISpot or flow cytometry tests. Such a criterion has an idea behind it of labeling patients who were earlier ill with COVID-19 as positive and those who were not as negative, in the same sense as positivity criteria for serology test developed by the manufacturer. The major limitation of this method arises from our inability to rule out earlier infection with SARS-Cov-2 by any other means except by using IgG levels.

#### ELISpot Positivity Criteria

For each ELISpot measurement, there are four main values provided in this study: number of spots (NoS) for M protein, NoS for N protein, NoS for S protein, and NoS for negative control. To build positivity criteria for each of the proteins, NoS for this protein and for negative control were used.

The commonly used positivity criteria in ELISpot are based on comparison of negative control with experiment and selecting some additive and multiplicative boundary relations between these two values. Such a method, however, is based on an underlying hypothesis that a value observed in a negative control is not a random value but rather results from some underlying sample characteristic which may affect both non-control and negative control values. In order to check this hypothesis and select the optimal positivity criteria, the following testing procedure was used.

We supposed that there exists a two-parametric threshold which separates positive and negative groups of patients which can be expressed as NoS_[protein] = a*NoS_[negative_control]+b. We then selected a set of fixed ‘a’ <OUT_OF> [0;0.5;1;2], and for each ‘a_i’ from this set we tested the all possible ‘b’ values to calculate the optimal false positive rate (FPR) and true positive rate (TPR) and build a ROC curve. For such a ROC curve, the area under the curve AUC_i was calculated, and optimal ‘b_i’ was selected (as ‘b_i’ which minimizes Euclidian distance to (FPR=0, TPR=1) point, FPR^2+(1-TPR)^2). We then compared AUC_i for different a_i (see Supplementary Figure S8).

We found that, although for the N protein ROC AUC was maximized with a=0, for the M protein with a=0.5, and for the S protein with a=1, in all cases the differences between AUC for all ‘a’ values were minor. The resulting optimal TPR and FPR for each of the curves were nearly identical, and all models performed nearly identically. This arises from the fact that multiplication factor ‘a’ will come into play only for the larger values of NoS in the negative control, which are effectively absent from the dataset, and for smaller values only the additive part, ‘b’, is important. Thus, we decided to set ‘a’ equal to zero and to use positivity criteria which suppose independence of experimental values from negative control values. The optimal values of ‘b’ were 9, 4, and 13 for the M, N, and S proteins, respectively, meaning that samples with protein NoS above this threshold were labeled as positive, and samples with protein NoS below or equal to this threshold were labeled as negative for the corresponding protein.

For all proteins, the FPR fell in the range ∼0.15–0.2, and optimal TPR fell in the range of ∼0.8–0.9, which in a sense characterizes the ability of the positivity criteria to correctly distinguish patients who were earlier ill with COVID-19 from those who were not.

#### Flow Cytometry Positivity Criteria

For flow cytometry positivity criteria, an analysis similar to the ELISpot data one was used. Fractions of CD4^+^ or CD8^+^ T cells expressing IFNγ only, IL2 only, or both cytokines out of the total number of CD4^+^ or CD8^+^ T cells, respectively, were calculated in experiment and negative control; we also additionally calculated fractions of all CD4^+^, CD8^+^ cells expressing IFNγ and all CD4^+^, CD8^+^ cells expressing IL2. Thus, experimental value and negative control were paired for each type of cell fractions and were used in the search for the optimal threshold of Fraction_[experiment] = a*Fraction_[negative_control]+b type for ‘a’ <OUT_OF> [0;0.5;1;2].

However, there was a significant difference between the positivity analysis performed in ELISpot and that in the flow cytometry studies: since the study was multi-central and different devices were used, the positivity criteria were evaluated independently for each of the three study centers, labeled below as Center #1, #2 and #3.

Supplementary Table S1 contains information about the highest AUC achieved, and the corresponding ‘a’ value and optimal ‘b’ for this ‘a’ for each cell fraction and each center (see Supplementary Figure S9 for an example of analysis). The analysis of results allowed us to make the following conclusions:

- positivity criteria were different for different centers;
- in many cases, the maximum ROC AUC was achieved for non-zero ‘a’ and was higher than for ROC with *a*=0 (data not provided; see Supplementary Figure S9 for an example in the case of the fraction of CD4^+^ T cells expressing IFNγ only);
- for Center #2, the maximum ROC AUC was achieved for *a*=0, which signifies the independence between negative control and experiment, while for Centers #1 and #3 the maximum ROC AUC was always achieved for non-zero *a*;
- the ROC AUC for separation of two groups was the highest in Center #3, lower in Center #2, and lowest in Center #1 for all CD4^+^ cell fractions;
- among different CD8^+^ cell fractions, only the fraction of CD8^+^ T cells expressing IFNγ allowed us to distinguish between positive and negative groups, with AUC considerably lower if compared to CD4^+^ cell fractions.

As a result of these observations, in contrast with ELISpot positivity criteria selection, we did not select *a*=0 as a pre-set parameter but rather used optimal ‘a’ and ‘b’ resulting from the analysis. Criteria selected for each center and each cell fraction were afterwards used only for the data coming from this particular center. At the same time, it should be noted that for all cases ROC AUC for *a*=0 was comparable or equal to ROC AUC for optimal ‘a’, which means that cell fractions can be used as a quantitative metric of T cell immune response without additional corrections by negative control.

**Supplementary Table S1.**

| Cell Fraction | Center | optimal a | optimal b | ROC AUC<br>for optimal a |
| --- | --- | --- | --- | --- |
| **CD4<sup>+</sup>** |  |  |  |  |
| Fraction of CD4<sup>+</sup>IFNγ<sup>+</sup> out of all CD4<sup>+</sup> | Center # 1 | 1 | 1.7E-04 | 0.631 |
|  | Center # 2 | 0 | 4.9E-04 | 0.726 |
|  | Center # 3 | 1 | 1.7E-04 | 0.783 |
| Fraction of CD4<sup>+</sup>IL2<sup>+</sup> out of all CD4<sup>+</sup> | Center # 1 | 1 | 1.8E-04 | 0.677 |
|  | Center # 2 | 0 | 2.3E-04 | 0.732 |
|  | Center # 3 | 0.5 | 3.5E-04 | 0.854 |
| Fraction of CD4<sup>+</sup>IL2<sup>+</sup>IFNγ<sup>+</sup> out of all CD4<sup>+</sup> | Center # 1 | 2 | 8.7E-05 | 0.721 |
|  | Center # 2 | 0 | 8.5E-05 | 0.732 |
|  | Center # 3 | 0.5 | 1.6E-04 | 0.915 |
| Fraction of all CD4<sup>+</sup>IFNγ<sup>+</sup> out of all CD4<sup>+</sup> | Center # 1 | 2 | 2.0E-04 | 0.649 |
|  | Center # 2 | 0 | 5.3E-04 | 0.740 |
|  | Center # 3 | 1 | 5.3E-04 | 0.848 |
| Fraction of all CD4<sup>+</sup>IL2<sup>+</sup> out of all CD4<sup>+</sup> | Center # 1 | 2 | 1.8E-04 | 0.704 |
|  | Center # 2 | 0 | 2.8E-04 | 0.745 |
|  | Center # 3 | 0.5 | 6.0E-04 | 0.881 |
| Fraction of all activated CD4<sup>+</sup> | Center # 1 | 1 | 8.1E-04 | 0.676 |
|  | Center # 2 | 0 | 6.7E-04 | 0.757 |
|  | Center # 3 | 0.5 | 8.5E-04 | 0.847 |
| **CD8<sup>+</sup>** |  |  |  |  |
| Fraction of CD8<sup>+</sup>IFNγ<sup>+</sup> out of all CD8<sup>+</sup> | Center # 1 | 2 | -1.3E-03 | 0.581 |
|  | Center # 2 | 0 | 5.6E-04 | 0.689 |
|  | Center # 3 | 0.5 | 2.3E-04 | 0.648 |
| Fraction of CD8<sup>+</sup>IL2<sup>+</sup> out of all CD8<sup>+</sup> | Center # 1 | 2 | 1.5E-07 | 0.525 |
|  | Center # 2 | 0 | 3.2E-05 | 0.535 |
|  | Center # 3 | 2 | -9.5E-05 | 0.546 |
| Fraction of CD8<sup>+</sup>IL2<sup>+</sup>IFNγ<sup>+</sup> out of all CD8<sup>+</sup> | Center # 1 | 2 | 0.0E+00 | 0.574 |
|  | Center # 2 | 0 | 3.4E-05 | 0.520 |
|  | Center # 3 | 0 | 2.8E-05 | 0.614 |
| Fraction of all CD8<sup>+</sup>IFNγ<sup>+</sup> out of all CD8<sup>+</sup> | Center # 1 | 2 | -1.0E-03 | 0.581 |
|  | Center # 2 | 0 | 8.4E-04 | 0.683 |
|  | Center # 3 | 0.5 | 2.5E-04 | 0.647 |
| Fraction of all CD8<sup>+</sup>IL2<sup>+</sup> out of all CD8<sup>+</sup> | Center # 1 | 2 | 1.7E-05 | 0.537 |
|  | Center # 2 | 0 | 6.6E-05 | 0.549 |
|  | Center # 3 | 1 | 2.6E-05 | 0.588 |
| Fraction of all activated CD8<sup>+</sup> | Center # 1 | 2 | -7.7E-04 | 0.591 |
|  | Center # 2 | 0 | 8.9E-04 | 0.690 |
|  | Center # 3 | 0.5 | 2.9E-04 | 0.634 |

### Supplementary Section 2: Comparison of different immune response metrics

For metrics of T cell response evaluated with ELISpot and flow cytometry and for IgG/IgM measures by Mindray assay, the Spearman correlation matrix was calculated as shown in Supplementary Figure S4.

#### NoS for M, N, and S protein correlations

For all three pairwise comparisons of M, N, and S protein NoS, a high Spearman correlation coefficient was observed (M vs. N, 0.69; S vs. N, 0.69; S vs. M, 0.75; see also Supplementary Figure S4). NoS for the S protein was statistically significantly higher than NoS for the M and N proteins, and NoS for the M protein was higher than NoS for the N protein, although the values were very close (Wilcoxon paired test *p*-value < 1e-19 for all comparisons). It is most likely that these results originate from the differences in sizes of these proteins, since the S protein is the largest among them, while the M and N protein sizes are comparable to one another and both are smaller than the S.

#### Flow Cytometry: CD4+ and CD8+ cell fraction correlations

For all three pairwise comparisons of the fractions of CD4^+^ cells expressing IFNγ only, IL2 only, or both cytokines, statistically significant Spearman correlation coefficients were observed (IL2^+^ vs. IFNγ^+^, 0.41; IFNγ^+^ vs. IL2^+^IFNγ^+^, 0.58; IL2^+^ vs. IL2^+^IFNγ,: 0.75; see also Supplementary Figure S4). The fraction of CD4^+^ cells expressing IL2 only was significantly lower than the fraction of cells expressing IFNγ only (Wilcoxon paired test *p*-value < 0.002), and the fraction of cells expressing both cytokines was lower than either of fractions of cells expressing one cytokine only (Wilcoxon paired test *p*-value < 1e-100).

Additionally, the fraction of CD4^+^ cells expressing IFNγ only out of the total number of CD4^+^ cells was compared with the fraction of CD8^+^ cells expressing IFNγ only out of the total number of CD8^+^ cells, and a statistically significant Spearman correlation coefficient of 0.53 was observed. Similarly, the fraction of CD4^+^ cells expressing IL2 only out of the total number of CD4^+^ cells was correlated with the fraction of CD8^+^ cells expressing IL2 only out of total number of CD8^+^ cells, with a Spearman correlation coefficient of 0.47 (see also Supplementary Figure S4).

#### Correlation between ELISpot and flow cytometry

In order to perform additional comparisons between ELISpot and flow cytometry, we computed the fraction of all CD4^+^/CD8^+^ cells expressing IFNγ (cells also expressing IL2 included) and summed NoS for M, N, and S. No correlation was observed between any ELISpot NoS and any fractions of CD8^+^ cells. On the contrary, NoS for all proteins and their combination was significantly correlated with all CD4^+^ cell fractions, with a Spearman correlation coefficient in a range from 0.38 to 0. 56 for different comparison pairs (see also Supplementary Figure S4).

#### Correlation between IgG antibody levels and T cell immune response metrics

A Spearman correlation of ∼0.6 was observed between IgG and ELISpot NoS for each of three proteins. Compared with flow cytometry results, IgG antibody levels were most strongly correlated with the fraction of CD4^+^ cells expressing both IFNγ and IL2 out of the total number of CD4^+^ cells (Spearman correlation coefficient 0.52), followed by those expressing the IL2-only fraction (Spearman correlation coefficient 0.45) and the IFNγ-only fraction (Spearman correlation coefficient 0.34). None of the CD8^+^ cell fractions demonstrated a high correlation with IgG antibody levels (see also Supplementary Figure S4).

We additionally studied whether the observed correlation is only a result of the existence of two main groups of patients, one with very low responses for both IgG and T cell immunity and another with high responses for both cases, or whether there is an intermediate group of patients with both an antibody response and an intermediate T cell response. In order to do that, we analyzed the distribution of S NoS and of the fraction of CD4^+^ cells expressing both IFNγ and IL2 out of the total number of CD4^+^ cells as a function of IgG levels split by deciles, using ridgeline plots (Supplementary Figure S10). For the fraction of CD4^+^ cells expressing both IFNγ and IL2 out of the total number of CD4^+^ cells, there is a smooth transition not only with average levels but also with the whole distribution moving towards higher values as IgG levels were increased (similar results were observed for all other CD4^+^ cell fractions; not given in the paper). On the contrary, for S NoS there was no transition, but the density distribution for intermediate levels can be interpreted as a mixture of distributions observed for high and low IgG levels (similar results were observed for M and N NoS; not given in the paper). It should be noted that results provided for ELISpot NoS should be viewed with some doubt, since the distribution of NoS is inherently not normal, and ridgeline plots employ kernel-density estimates using Gaussian kernels.

### Supplementary Section 3: Kaplan-Meyer Curves

We observed all the individuals included in the study using the Moscow State COVID-19 registry, which includes information about patient illness and vaccination. In order to perform an analysis using Kaplan-Meyer curves, we excluded from the analysis individuals with pre-inclusion clinically confirmed COVID-19, as well as those who were included in the clinical trial of vaccine or who had been vaccinated before the inclusion. Next, we excluded all the patients who were added into the registry as ill within five days after inclusion in the study in order to exclude patients who might already have been ill at the moment of inclusion and blood collection. All the remaining participants were then considered as eligible for post-inclusion observation. Thus, by the end of observation at the beginning of April, 2021, among the 4 027 participants who were eligible for the post-inclusion observation, a total of 496 post-inclusion cases of COVID-19 were registered.

For these participants, we employed the Kaplan–Meier estimator. The date of the patient’s illness according to the Moscow State COVID-19 registry was considered to be the date of the ‘event’, while the date of vaccination or the date of last observation, whichever came first, was considered to be the right-censoring date. After that, the classical Kaplan-Meier estimator was used to study the survival function in different subgroups of patients.

### Supplementary Section 4: Correlation between different serological tests and virus-neutralizing activity

Within the cohort we selected a group of individuals whose IgG titers, estimated using the automated analyzer (Mindray, China; here and below mentioned as ‘IgG (Mindray)’), were uniformly represented. Because of the limited sample volume, these individuals were additionally tested either for virus-neutralizing activity (VNA) and spike S-protein-specific IgG antibodies or for S- and nucleocapsid N-protein specific IgGs.

We first analyzed relations between S-protein–specific antibodies, N-protein–specific antibodies, and IgG (Mindray). We detected a strong correlation between S- and N-protein– specific antibodies. We manually split all the individuals into four groups: a negative group, a group with correlated response, and groups with either only S-protein or only N-protein– specific antibodies (Supplementary Figure S2, panel A). The vast majority of the individuals with some response demonstrated antibodies against both S- and N-protein (76.5%), while S- and N-protein single-positive individuals represented only 8.4 and 15.1%, respectively. These data are in good agreement with the results of the ELISpot shown in Figure 3C.

We also found a strong correlation between automated analyzer IgG (Mindray) titers and both S-protein and N-protein–specific antibodies. These results are in accordance with the manufacturer’s description, since this serological test was developed to detect antibodies specific to the full length of SARS-CoV-2 N protein and receptor binding domain (RBD) portion of the spike SARS-CoV-2 protein (Supplementary Figure S2, panel A).

We additionally analyzed the connections between VNA, S-protein–specific antibodies, and IgG (Mindray). We showed that VNA correlated with both IgG (Mindray) titer and S-protein-specific antibody levels (Supplementary Figure S2, panel B). The highest VNA was detected in individuals with high IgG (Mindray) titer and high S-protein–specific antibody levels, making up the Q4 and Q5 quantiles of IgG (Mindray) (see Figure 5A and the corresponding section of the main text for description of quantiles). Particularly, we found a significant virus-neutralizing activity among individuals making up the Q3 quantile (with IgG (Mindray) values in a range 0.97–8.33 AU/mL). This finding probably explains the protection against SARS-CoV-2 infection observed for this quantile. However, there was no difference in VNA between Q1 and Q2 quantiles. It is still possible that Q2 possesses VNA that are lower than the minimal plasma dilution used in our study, and protectivity found for Q2 may be also explained by the presence of VNA in this group. Still, in this case VNA would be significantly lower in the Q2 quantile than in Q3.

---

**Supplementary Figure S1.**
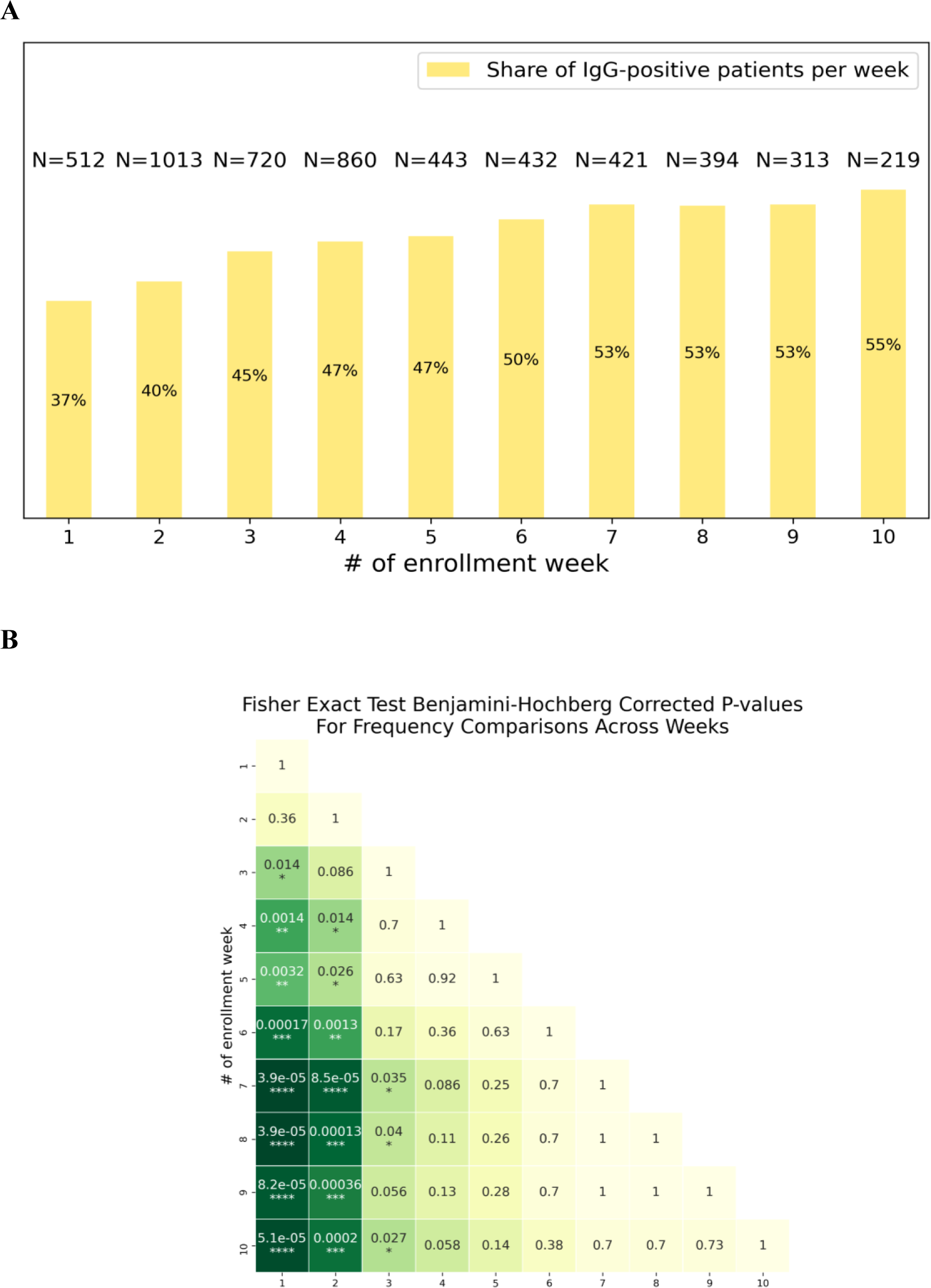
Changes in share of IgG-positive patients per enrollment week **(A):** Share of IgG-positive participants per enrollment week. Number of participants per week (N) is given above each bar. **(B):** Benjamini-Hochberg corrected p-values for pairwise comparison of share of IgG-positive patients per week of inclusion.

**Supplementary Figure S2.**
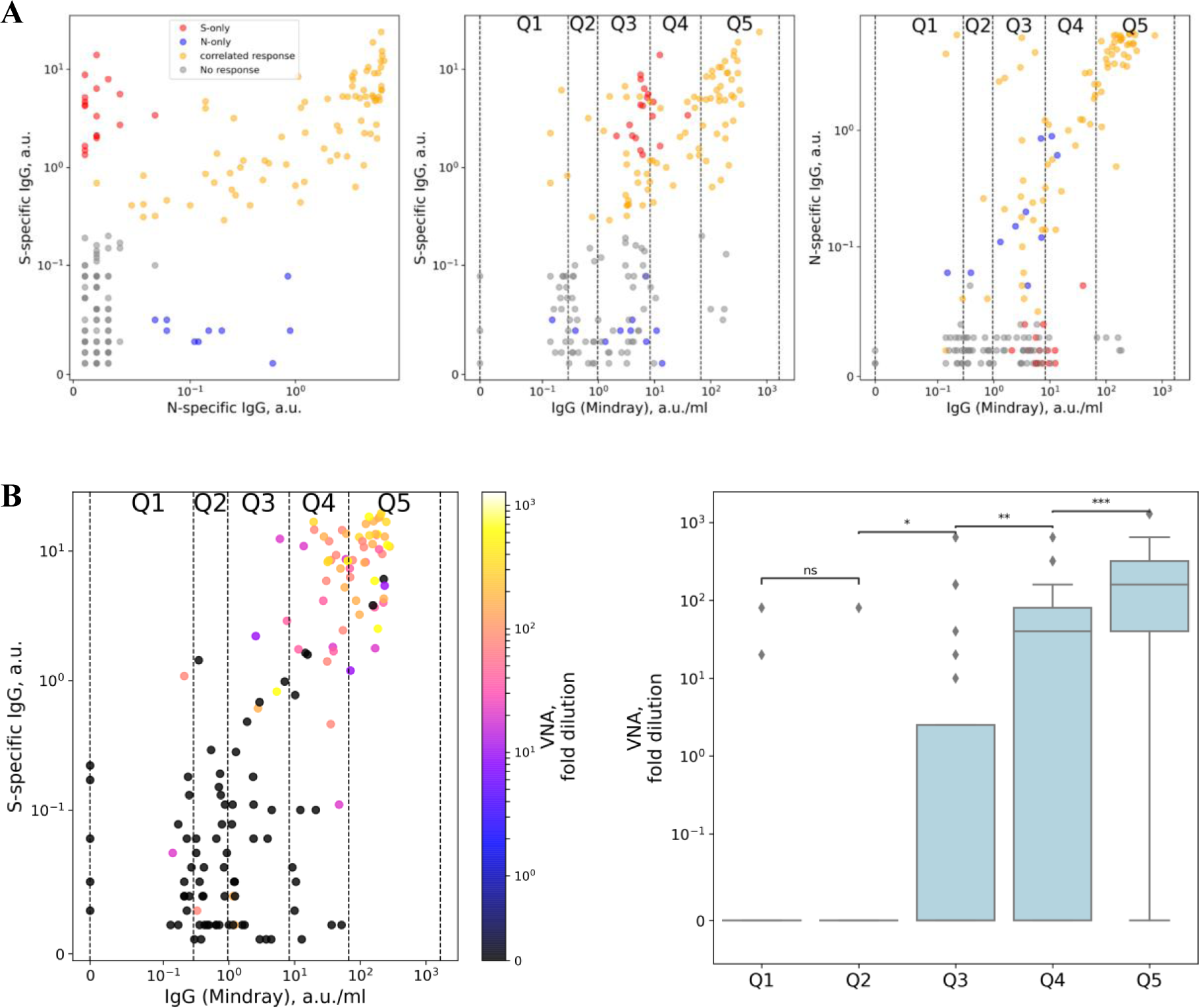
Correlation between different serological tests and virus-neutralizing activity. **(A)** Comparisons of S-protein–specific antibodies with N-protein–specific antibodies (left), S-protein specific antibodies with IgG (Mindray) (middle), and N-protein–specific antibodies with IgG (Mindray) (right). All samples were manually splitted into four groups: a group with no response (shown in gray), a group with correlated response (shown in yellow), and groups with either only S-protein–specific or only N-protein–specific antibodies (shown in red and blue, respectively). In the middle and right panels, quantiles for IgG (Mindray) as used in Figure 5 are marked as vertical lines and labeled from Q1 to Q5. **(B) Left:** Comparison of S-protein–specific antibodies with IgG (Mindray) colored with VNA measured in the same sample. Quantiles for IgG (Mindray) as used in Figure 5 are marked as vertical lines and labeled from Q1 to Q5. **Right:** Comparison of VNA in each quantile. Pairwise comparisons by Mann-Whitney test were performed for all neighboring pairs of quantiles.

**Supplementary Figure S3.**
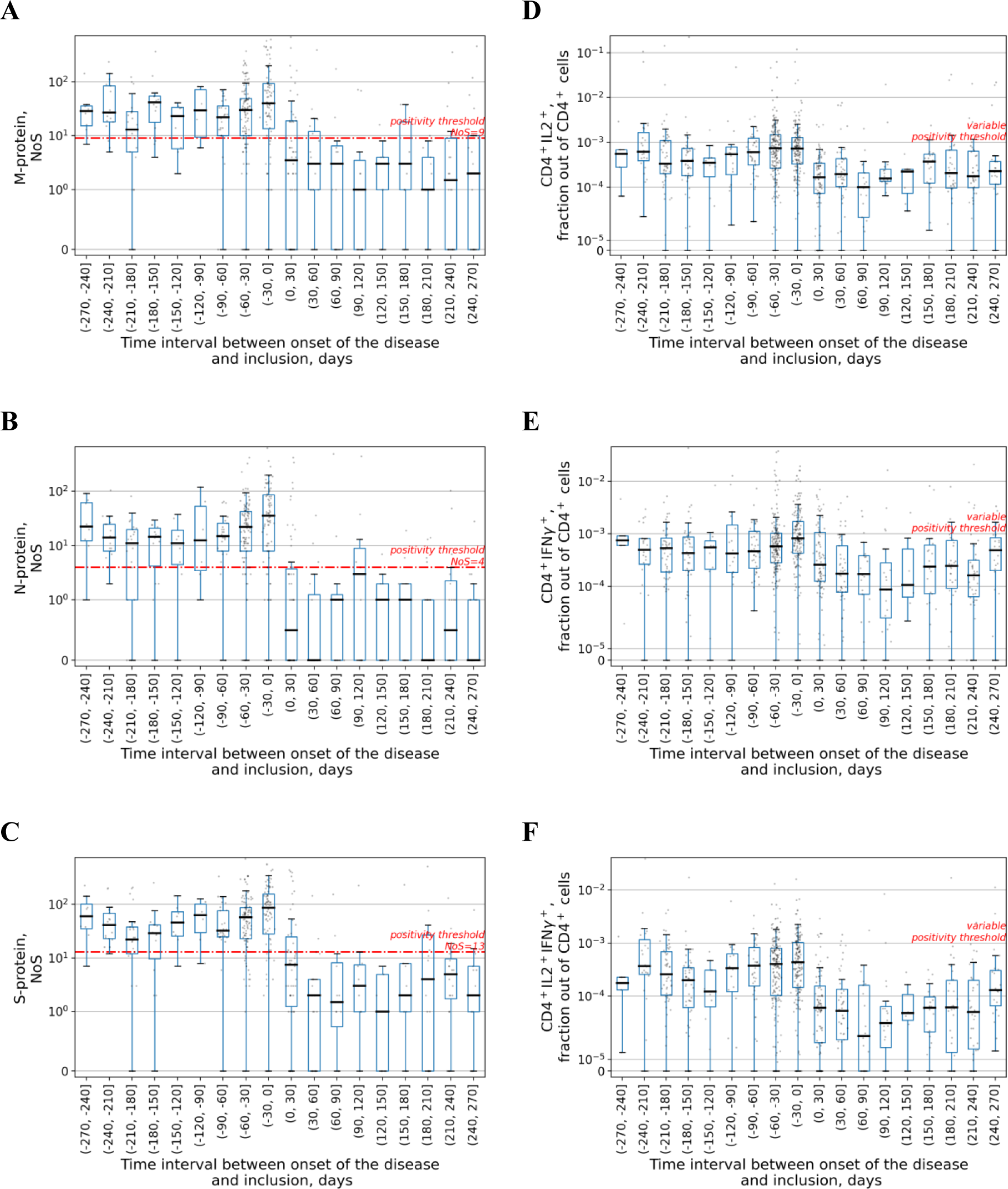
Temporal stability of all ELISpot and flow cytometry **(A),(B),(C):** Time dependence of the spots number for M, N, and S proteins in ELISpot assay, respectively. Each dot represents a single patient. Time is counted from the date of disease onset according to the Moscow State COVID-19 registry to the day of the inclusion in the study and thus serology testing. Time interval presented in each box-plot is 30 days. Red line represents a positivity threshold. **(D),(E),(F):** Time dependence of the percentage of CD4^+^ T cells expressing IL2, IFNγ, or both cytokines, respectively, in flow cytometry assay. Each dot represents a single patient. Time is counted from the date of disease onset according to the Moscow State COVID-19 registry to the day of the inclusion in the study and thus serology testing. Time interval presented in each boxplot is 30 days. Positivity threshold was variable (see Supplementary Materials for more details) and thus not given here.

**SupplementaryFigure S4.**
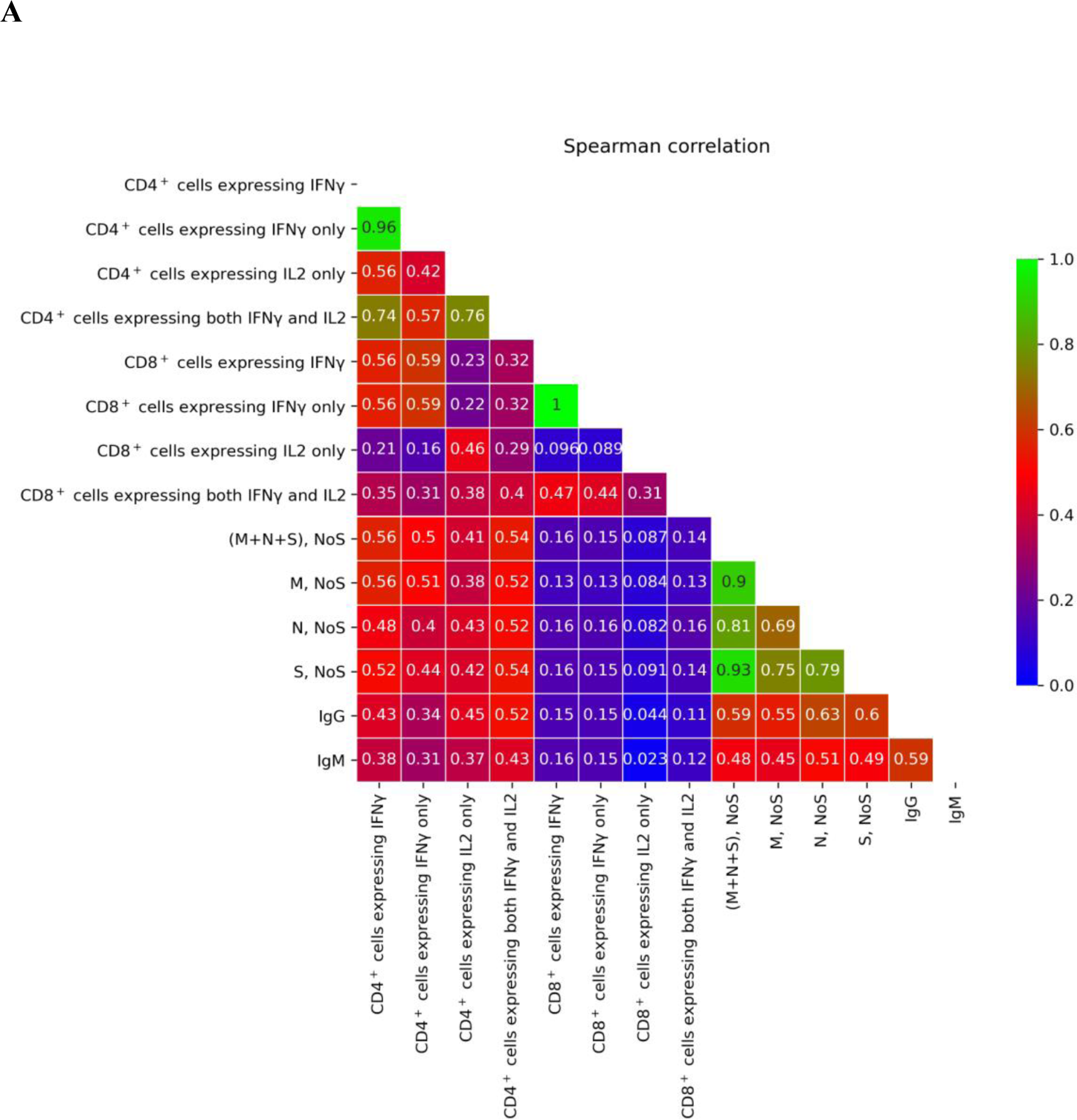
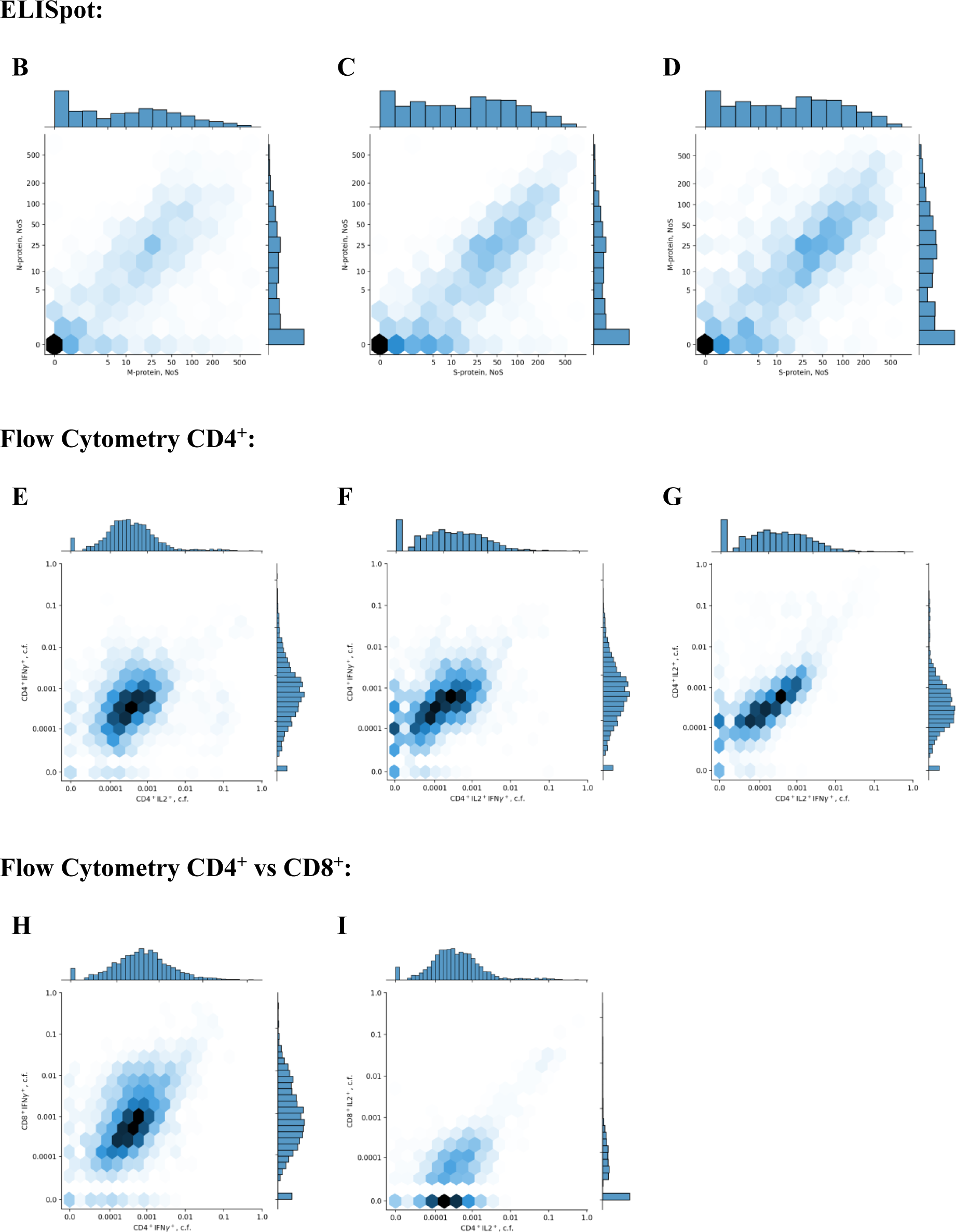
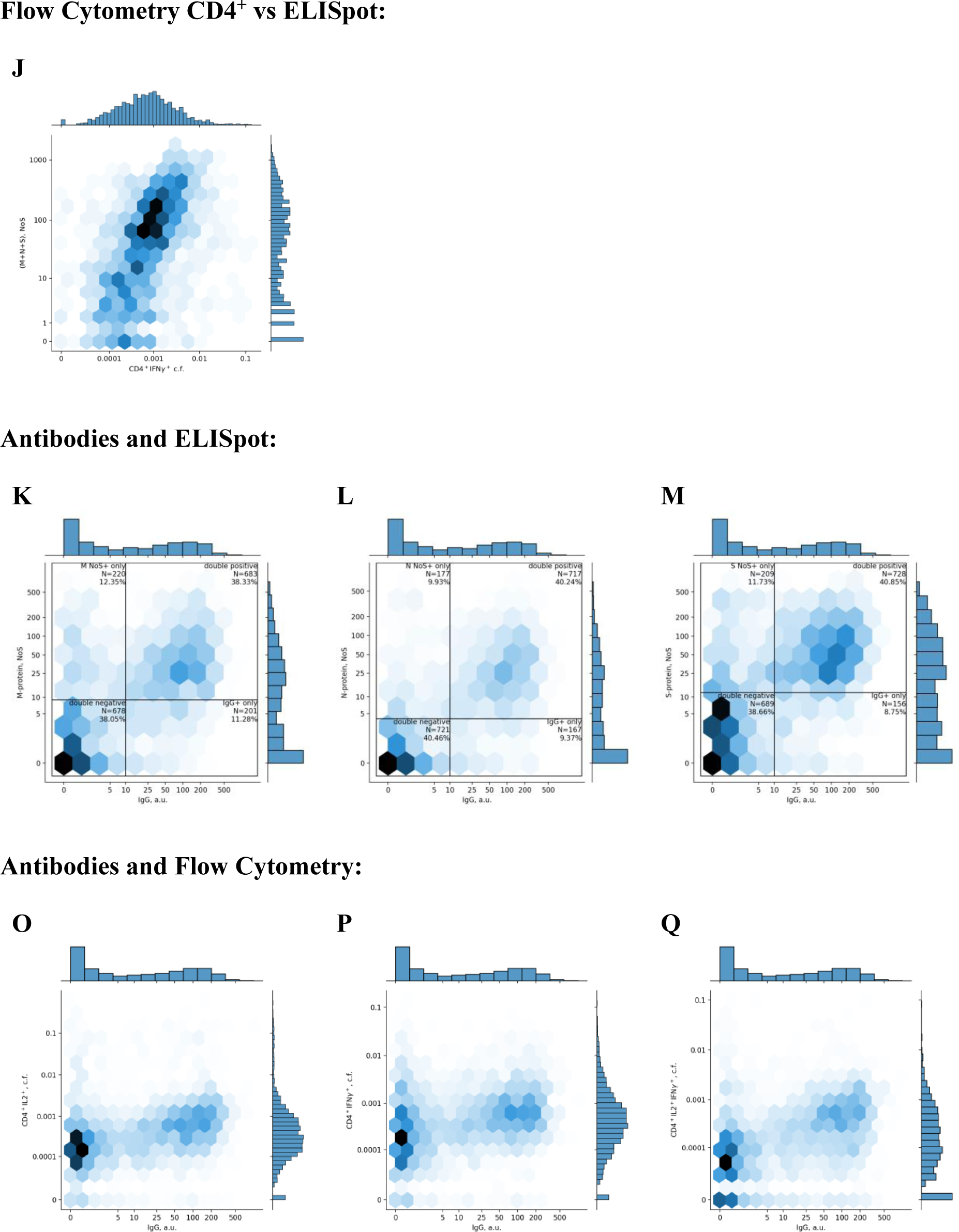
Comparison of different immune response metrics. **(A)** Spearman correlation coefficient matrix for different metrics of immune response, with low correlation colored in blue, middle in red, and high in green (no negative correlations were observed) **(B),(C),(D)** Density plots for pairwise comparison of NoS for different pairs of ELISpot proteins: M – S, S – N, and S – M, respectively. Each cell is colored according to the number of participants falling within this cell, with darker colors showing more participants. Histograms of value distribution are given along the corresponding axis. **(E),(F),(G)** Density plots for pairwise comparison of activated CD4^+^ cell fractions for different cytokines: IL2^+^ – IFNγ^+^, IL2^+^IFNγ^+^ – IFNγ^+^, and IL2^+^IFNγ^+^ – IL2^+^, respectively. Each cell is colored according to the number of participants falling within this cell, with darker colors showing more participants. Histograms of value distribution are given along the corresponding axis; ‘c.f.’ stands for ‘cell fraction’. **(H),(I)** Density plots for pairwise comparison of activated CD4^+^ and CD8^+^ cell fractions for different cytokines: CD4^+^IFNγ^+^ – CD8^+^IFNγ^+^ and CD4^+^IL2^+^ – CD8^+^IFNγ^+^, respectively. Each cell is colored according to the number of participants falling within this cell, with darker colors showing more participants. Histograms of value distribution are given along the corresponding axis; ‘c.f.’ stands for ‘cell fraction’. **(J)** Density plot for pairwise comparison of all activated CD4^+^ cells expressing IFNγ versus total number of spots for the M, N, and S proteins. Each cell is colored according to the number of participants falling within this cell, with darker colors showing more participants. Histograms of value distribution are given along the corresponding axis. **(K),(L),(M)** Density plots for pairwise comparison of levels of IgG and of NoS for different pairs of ELISpot proteins: M, N, and S, respectively. Each cell is colored according to the number of participants falling within this cell, with darker colors showing more participants. Histograms of value distribution are given along the corresponding axis. **(O),(P),(Q)** Density plots for pairwise comparison of levels of IgG and activated CD4^+^ cells fractions for different cytokines: IL2^+^, IFNγ^+^, and IL2^+^IFNγ^+^, respectively. Each cell is colored according to the number of participants falling within this cell, with darker colors showing more participants. Histograms of value distribution are given along the corresponding axis; ‘c.f.’ stands for ‘cell fraction

**Supplementary Figure S5.**
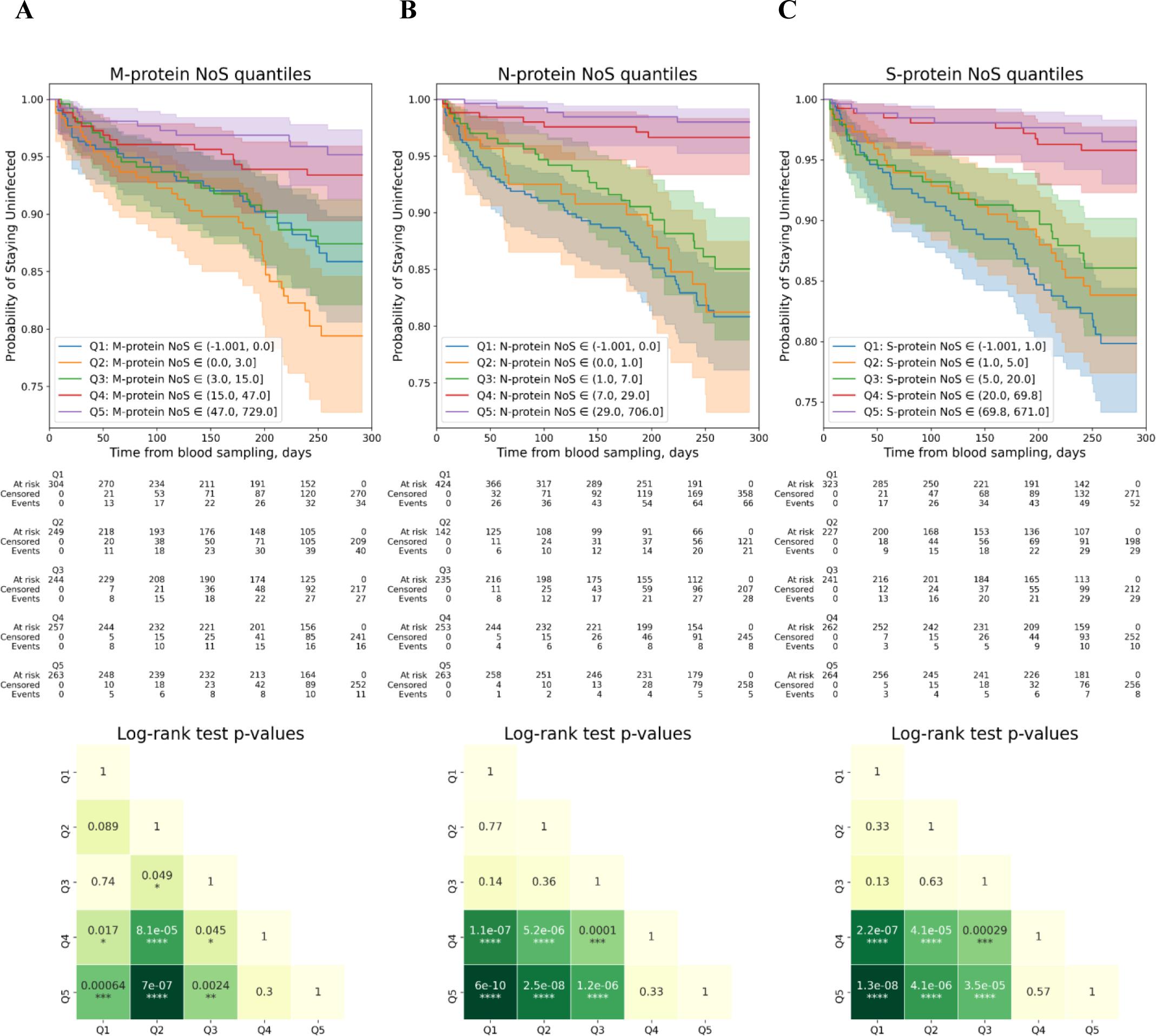
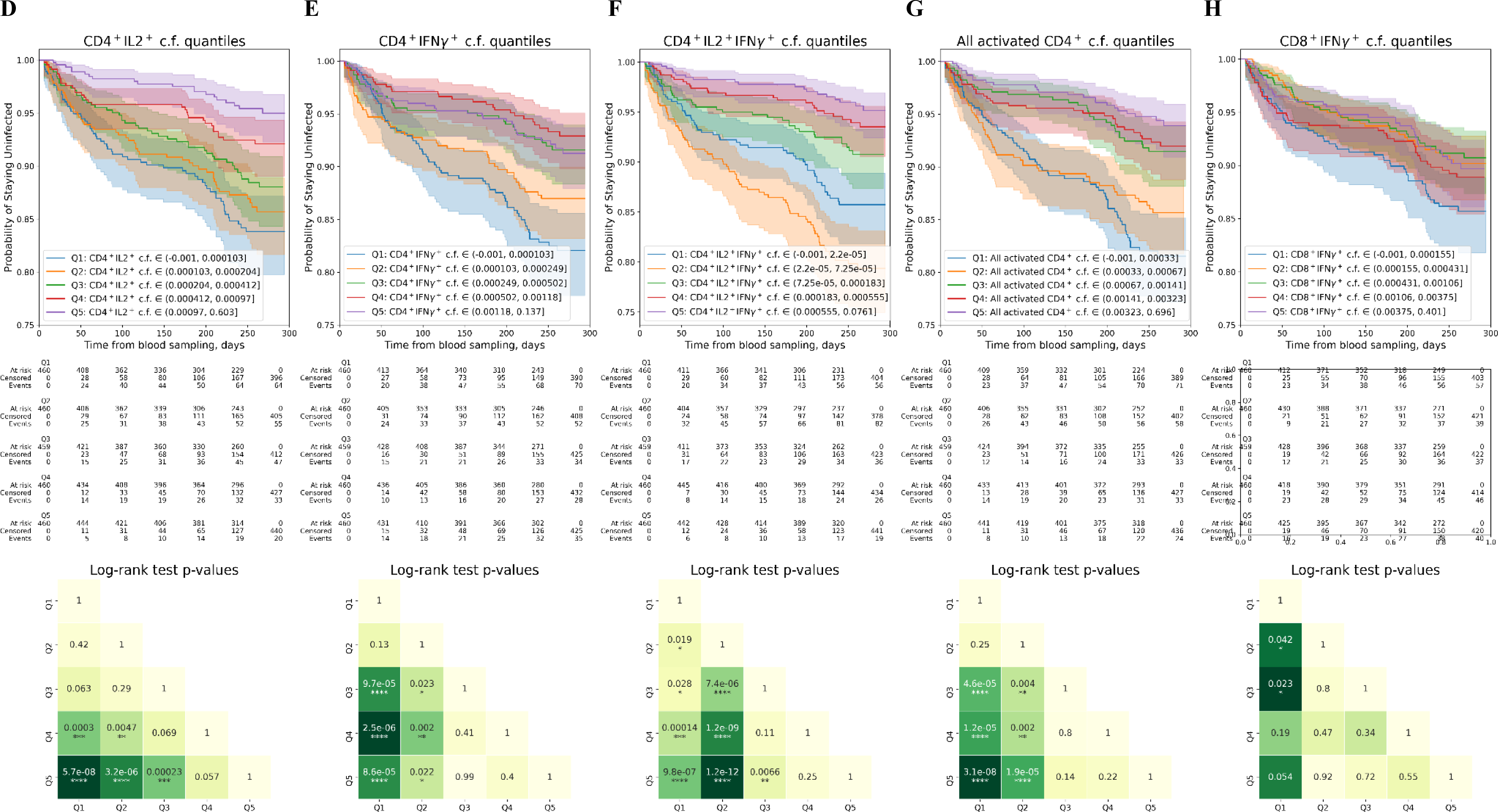
Evaluation of effects of T cell immunity on COVID-19 infection rates. **(A),(B),(C) Top**: Kaplan-Meyer curves for patients with different NoS for the M, N, and S proteins in ELISpot, respectively. The patients were split into five nearly equal groups by quantiles of NoS from Q1 to Q5, and a Kaplan-Meyer curve was built for each group (see Supplementary Materials for more details on Kaplan Meyer curves analysis). **Bottom**: log-rank test *p*-values for pairwise comparison of all five groups selected by quantiles. **(D),(E),(F),(G) Top**: Kaplan-Meyer curves for patients with different CD4^+^IL2^+^, CD4^+^IFNγ^+^, CD4^+^IL2^+^IFNγ^+^, and total activated CD4+ cell fractions out of all CD4^+^ T cells, respectively. The patients were split into five nearly equal groups by quantiles of cell fraction from Q1 to Q5, and a Kaplan-Meyer curve was built for each group (see Supplementary Materials for more details on Kaplan Meyer curves analysis); ‘c.f.’ stands for ‘cell fraction’. **Bottom**: log-rank test *p*-values for pairwise comparison of all five groups selected by quantiles. **(H) Top**: Kaplan-Meyer curves for patients with different CD8^+^IFNγ^+^ cell fractions out of all CD8^+^ cells. All the patients were split into five nearly equal groups by quantiles of cell fraction from Q1 to Q5, and a Kaplan-Meyer curve was built for each group (see Supplementary Materials for more details on Kaplan Meyer curves analysis); ‘c.f.’ stands for ‘cell fraction’. **Bottom**: log-rank test *p*-values for pairwise comparison of all 5 groups selected by quantiles

**Supplementary Figure S6.**
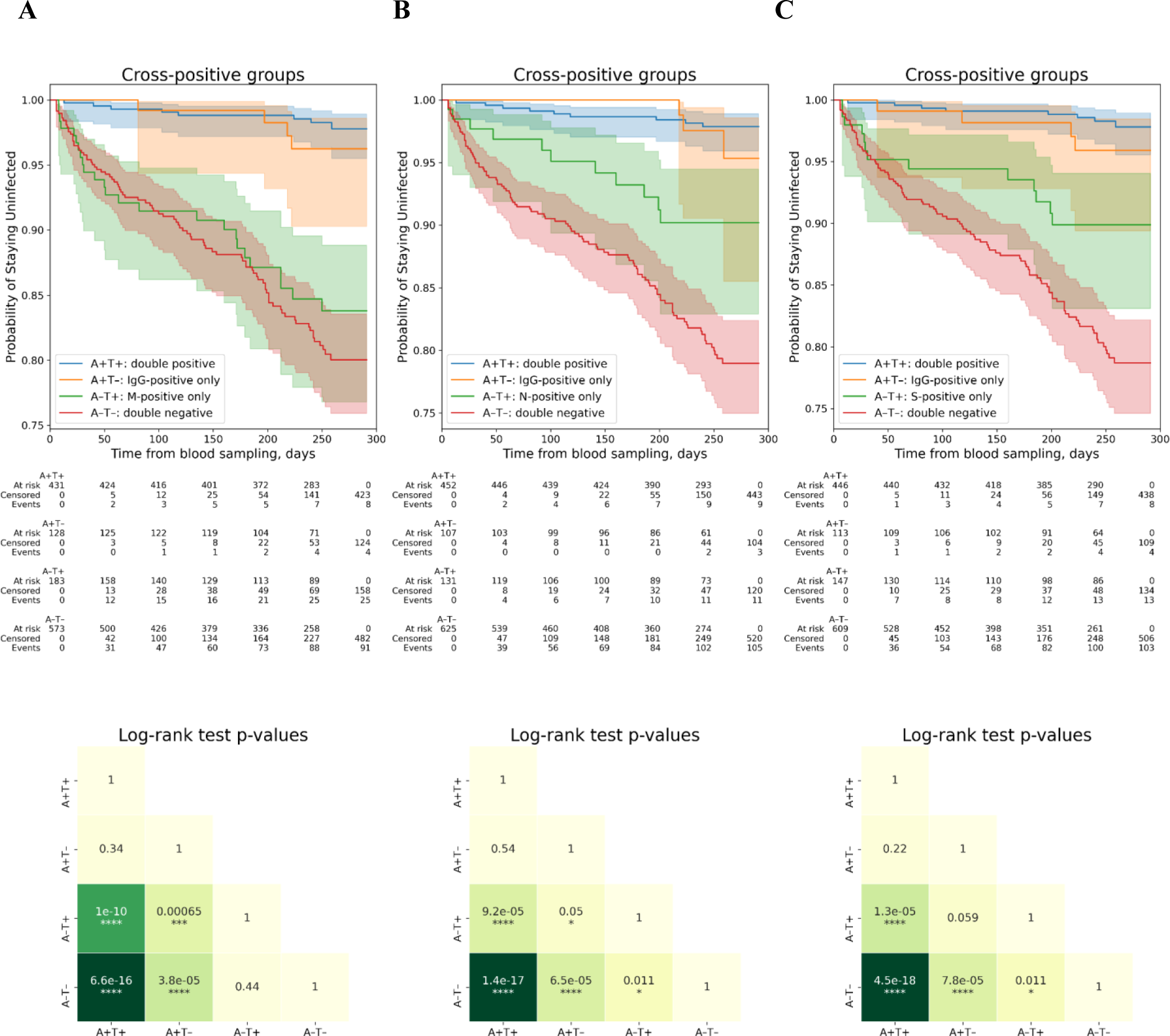
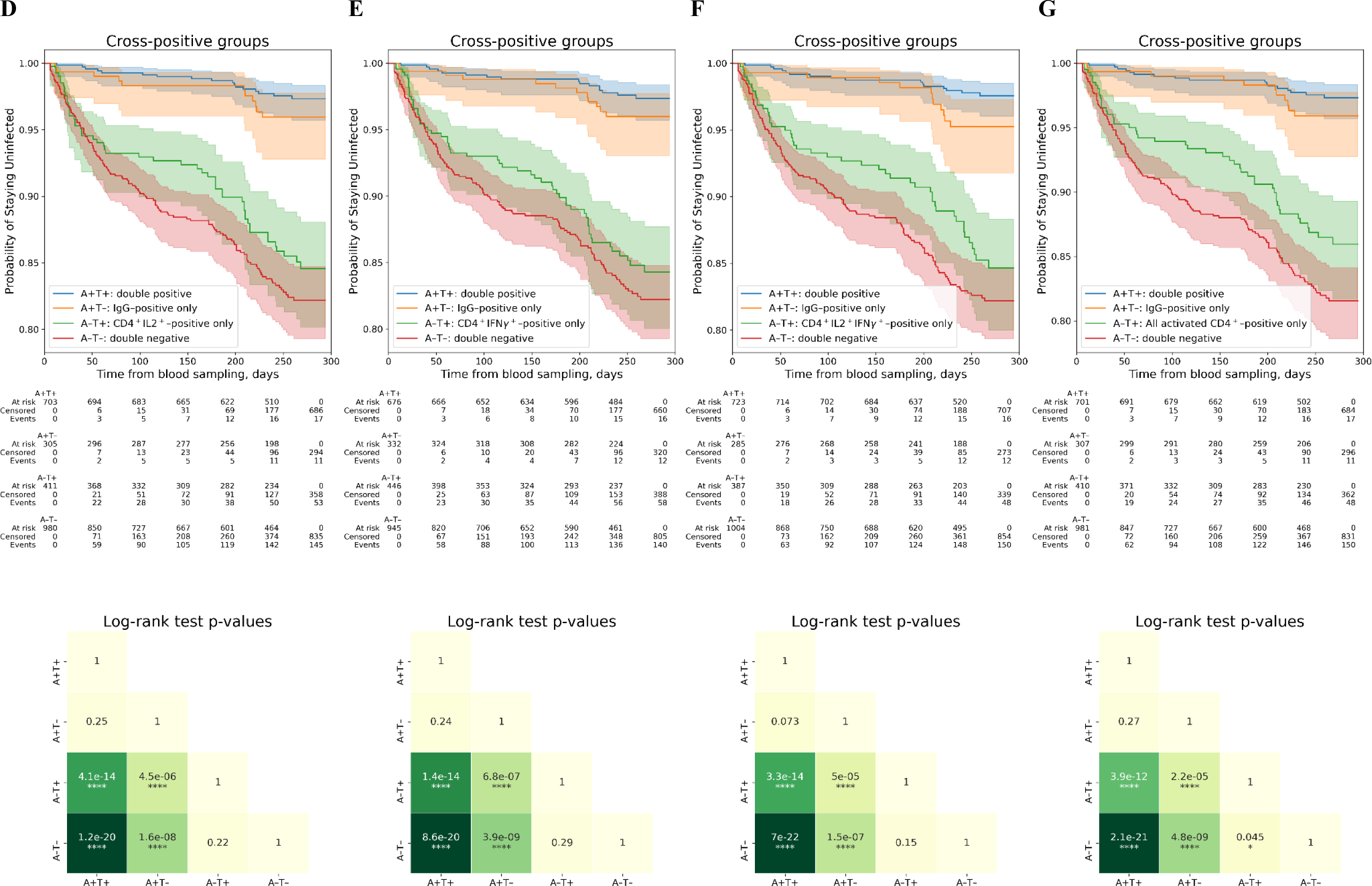
Evaluation of connection between effects of antibody immunity and T cell immunity on COVID-19 infection rates. **(A),(B),(C) Left:** Kaplan-Meyer curves for patients with different positivity by IgG levels and by NoS for the M, N, and S proteins, respectively. The participants were split into four groups: positive only by antibodies (A+T-), positive only by NoS (A-T+), double-positive (A+T+), and double-negative (A-T-), with the positivity criteria discussed above used for the evaluation, and a Kaplan-Meyer curve was built for each group (see Supplementary Materials for more details on Kaplan Meyer curves analysis). **Right:** log-rank test *p*-values for pairwise comparison of all four groups selected by positivity. **(D),(E),(F),(G) Left:** Kaplan-Meyer curves for patients with different positivity by IgG levels and by CD4^+^IL2^+^, CD4^+^IFNγ^+^, CD4^+^IL2^+^IFNγ^+^, and total activated CD4+ cell fractions out of all CD4^+^ T cells, respectively.. All the participants were split into four groups: positive only by antibodies (A+T-), positive only by cell fraction (A-T+), double-positive (A+T+), and double-negative (A-T-), with the positivity criteria discussed above used for the evaluation, and a Kaplan-Meyer curve was built for each group (see Supplementary Materials for more details on Kaplan Meyer curves analysis). **Right:** log-rank test *p*-values for pairwise comparison of all four groups selected by positivity.

**Supplementary Figure S7.**
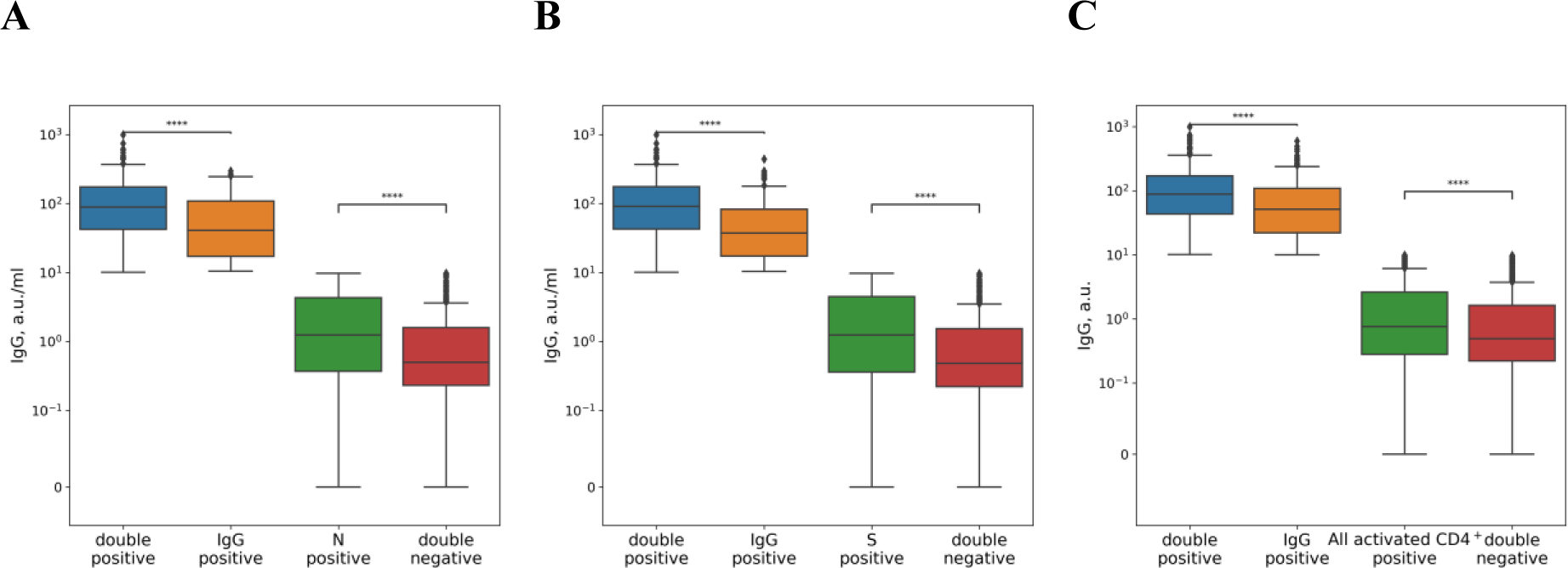
Comparison of IgG levels in different positivity groups. **(A),(B)** IgG levels for patients with different positivity by IgG levels and by N- and S-protein NoS in ELISpot, respectively. Comparison of groups with the same IgG positivity was performed by Mann-Whitney test and results are presented in the plot. **(C)** IgG levels for patients with different positivity by IgG levels and by total activated CD4^+^ T cell fractions out of all CD4^+^ cells in flow cytometry. Comparison of groups with the same IgG positivity was performed by Mann-Whitney test, and results are presented in the plot.

**Supplementary Figure S8.**
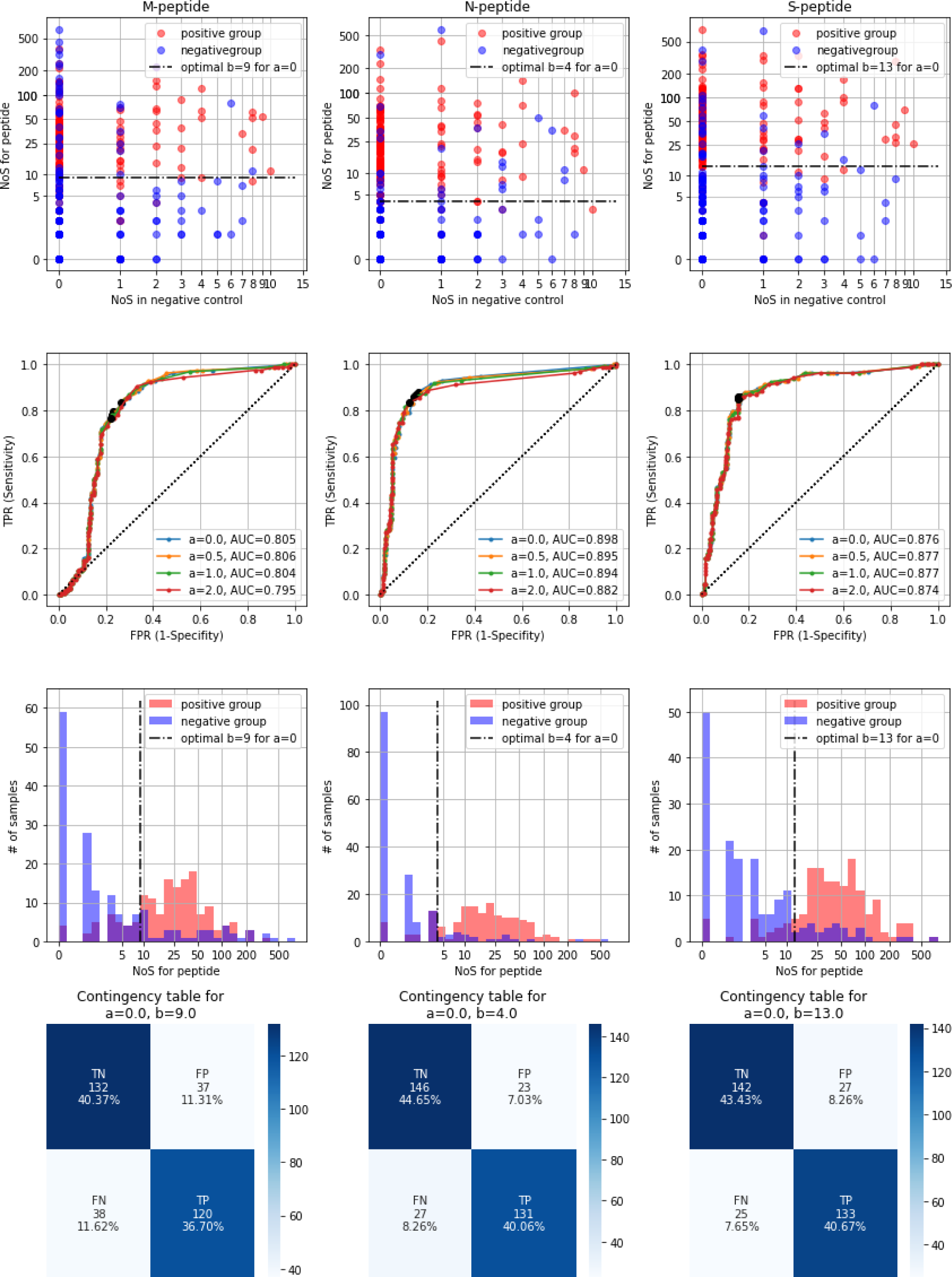
Selection of optimal positivity criteria for ELISpot based on patient groups. **Left column** shows results for M-protein; **middle,** for N-protein; **right**, for S-protein. **Top row:** scatter plots for connection between negative control and stimulated sample; one dot corresponds to one patient. Red dots are used to show patients from the positive group; blue dots are used to show patients from the negative group. The dotted black line shows optimal separation criteria selecting based on ROC curve analysis with *a*=0. **Second row from the top:** ROC curves for linear separation rule NoS_[protein] > a*NoS_[negative_control]+b with ‘a’ out of [0;0.5;1;2] and variable ‘b’. Black dots show the optimal point for each ROC curve (defined as the point of the curve closest to FPR=0, TPR=1). **Third row from top:** histograms showing distribution of NoS in experiment in positive group and negative group. **Bottom row:** contingency table for optimal rule selected on the basis of ROC curves for *a*=0 with ‘b’ above the table. ‘TN’ stands for ‘True Negative’, ‘TP’ for ‘True Positive’, ‘FN’ for ‘False Negative’, ‘FP’ for ‘False Positive’, with positive and negative group labels treated as true labels ‘1’ and ‘0’, respectively, and optimal rule-based labels used as predicted labels.

**Supplementary Figure S9.**
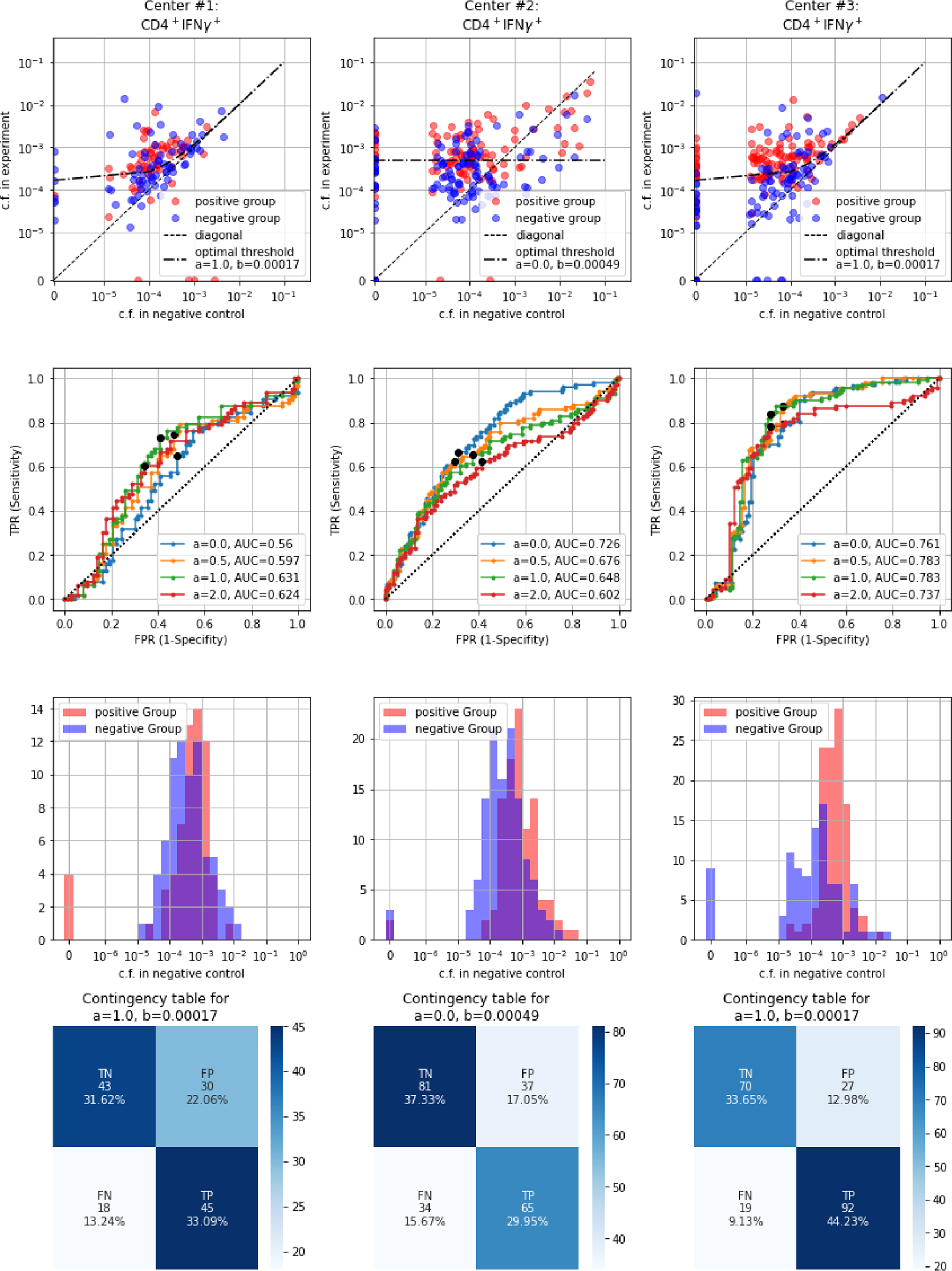
Selection of optimal positivity criteria for flow cytometry cell fractions based on patient groups. **Left column** shows results for the fraction of CD4^+^IFNγ^+^ cells out of all CD4^+^ cells in Center 1; **middle**, in Center 2; **right**, in Center 3. **Top row:** scatter plots for connection between negative control and stimulated sample; one dot corresponds to one patient. Red dots are used to show patients from the positive group; blue dots are used to show patients from the negative group. The dotted black line shows optimal separation criteria selected on the basis of ROC curve analysis. **Second row from the top:** ROC curves for linear separation rule Fraction_[experiment] > a*Fraction_[negative_control]+b with ‘a’ out of [0;0.5;1;2] and variable ‘b’. Black dots show the optimal point for each ROC curve (defined as the point of the curve closest to FPR=0, TPR=1). **Third row from top:** histograms showing distribution of cell fractions in experiment in positive group and negative group. **Bottom row:** contingency tables for optimal rule selected based on ROC curves with ‘a’ and ‘b’ above the table. ‘TN’ stands for ‘True Negative’, ‘TP’ for ‘True Positive’, ‘FN’ for ‘False Negative’, ‘FP’ for ‘False Positive’, with positive and negative group labels treated as true labels ‘1’ and ‘0’, respectively, and optimal rule-based labels used as predicted labels. In all panels, ‘c.f.’ stands for ‘cell fraction’

**Supplementary Figure S10.**
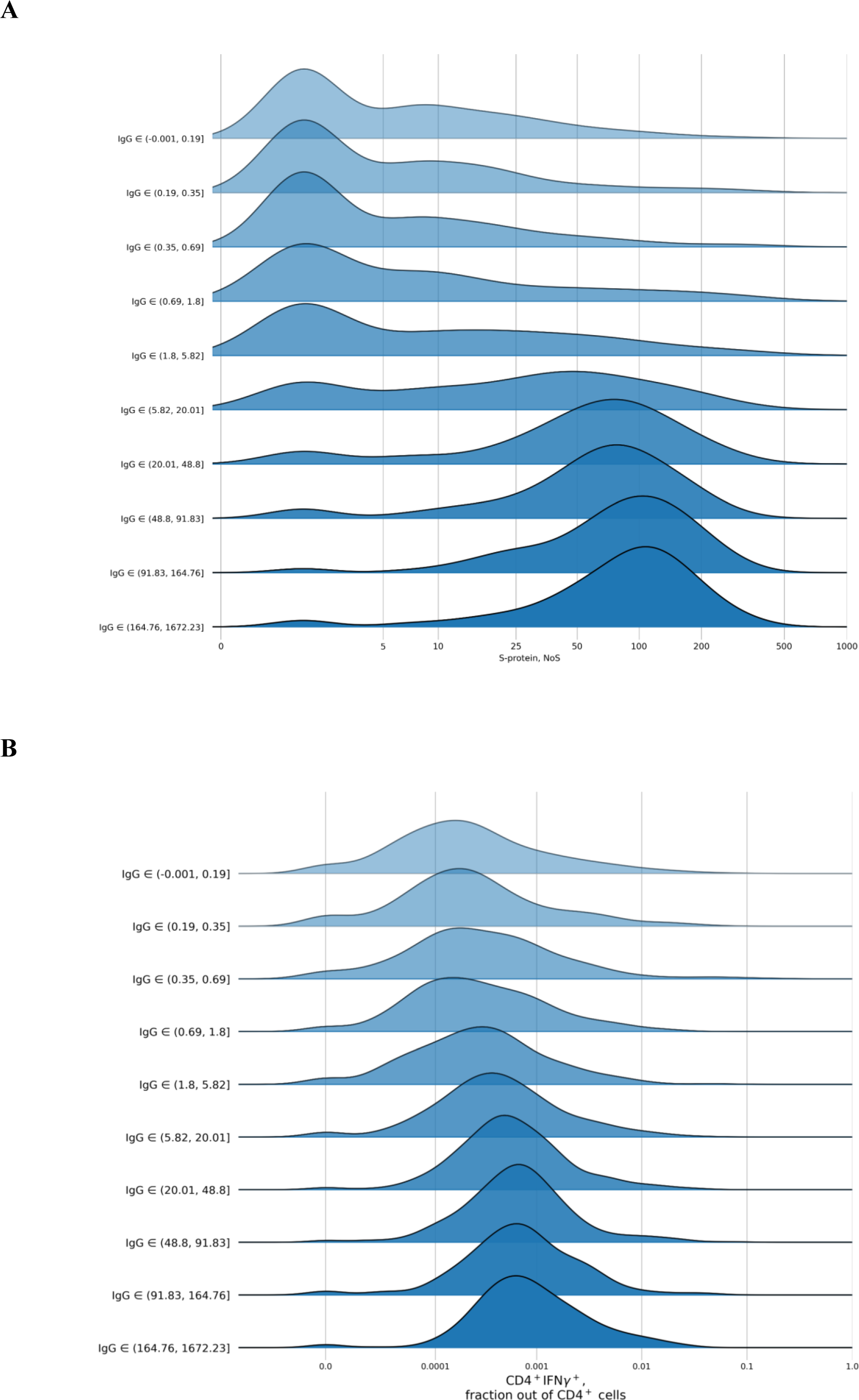
Ridgeline plots for T cell immunity metrics as a function of antibody levels. **(A), (B)** Distribution per decile of IgG of ELISpot S NoS and flow cytometry CD4^+^ cells expressing IFNγ out of the total number of CD4^+^ cells, respectively. For patients from each IgG decile, the plot shows the kernel-density estimate for the corresponding T cell immunity metric obtained using the Gaussian kernel.

